# Genetic variants associated with cross-disorder and disorder-specific risk for psychiatric disorders are enriched at epigenetically active sites in peripheral lymphoid cells

**DOI:** 10.1101/2021.08.04.21261606

**Authors:** Mary-Ellen Lynall, Blagoje Soskic, James Hayhurst, Jeremy Schwartzentruber, Daniel F. Levey, Gita A. Pathak, Renato Polimanti, Joel Gelernter, Murray B. Stein, Gosia Trynka, Menna R. Clatworthy, Ed Bullmore

## Abstract

Multiple psychiatric disorders have been associated with abnormalities in both the innate and adaptive immune systems. The role of these abnormalities in pathogenesis, and whether they are driven by psychiatric risk variants, remains unclear. We tested for enrichment of GWAS variants associated with multiple psychiatric disorders (cross-disorder or trans-diagnostic risk), or 5 specific disorders (cis-diagnostic risk), in regulatory elements in immune cells. We used three independent epigenetic datasets representing multiple organ systems and immune cell subsets. Trans-diagnostic and cis-diagnostic risk variants (for schizophrenia and depression) were enriched at epigenetically active sites in brain tissues and in lymphoid cells (T, B and NK cells), especially stimulated CD4^+^ T cells. There was no evidence for enrichment of either trans-risk or cis-risk variants for schizophrenia or depression in myeloid cells. This suggests a model where stimuli (e.g. infection) activate T cells to unmask the effects of psychiatric risk variants, contributing to the pathogenesis of mental health disorders.

## Introduction

Clinical diagnosis for mental health disorders is comprised of multiple, categorically distinct clinical syndromes such as schizophrenia, major depressive disorder (MDD), and bipolar disorder. However, symptoms overlap between some different psychiatric diagnoses, and comparative investigations of psychiatric disorders have revealed both shared and specific genetic ^1-3^ and environmental risk factors ^4,5^, and brain transcriptomic profiles ^6^. These and other data support a general predisposition to psychopathology or ‘p factor’ which captures an individual’s likelihood of developing any psychiatric disorder ^7^. Thus, some genetic and environmental risks operate trans-diagnostically across multiple psychiatric syndromes, rather than being cis-diagnostically aligned to a specific syndrome, as would be expected if each disorder was a biologically discrete disease entity.

Immune system abnormalities have been observed in case-control studies of many psychiatric disorders, including schizophrenia ^8,9^, MDD ^10^, bipolar disorder ^11^, autism spectrum disorder (ASD) ^12^, and attention deficit hyperactivity disorder (ADHD) ^13^. Among the most consistently reported findings, across multiple disorders, are increased C-reactive protein (CRP) ^14^, increased pro-inflammatory cytokines ^14,15^, increased white blood cell counts in both myeloid and lymphoid lineages ^16-22^, and inflammasome activation ^23-25^. Moreover, environmental exposures that elicit an immune response are risk factors for multiple psychiatric disorders, including *in utero* or parental infections ^26,27^, childhood and adult infections ^28-31^, childhood adversity ^32^, and acute or chronic stress ^33^. On this basis, it is conceivable that the immune system could be implicated in the pathogenesis of psychiatric disorders; but the direct evidence for a causal role of immune mechanisms is limited. Longitudinal studies have shown that immune dysregulation can be detected prior to onset of psychiatric disorder ^34^; but this could reflect the coincident effects of risk factors for psychiatric disorder, such as high body mass index (BMI), on both the immune system and the brain. Since germline genetic variants cannot be the consequence of disease, sequence variation associated with a disorder (or disorders) could shed light on the immune processes or cells likely to cause mental health symptoms. There is already some genetic evidence that psychiatric risk is mediated by the immune system. Polygenic risk scores (PRS) for depression, bipolar disorder and schizophrenia are associated with increased lymphocyte counts ^35^. Immune and psychiatric disorders are genetically correlated ^36,37^. Pathway analysis of genes trans-diagnostically associated with schizophrenia, bipolar disorder and MDD implicated neuronal, histone and immune pathways ^38^; although a larger trans-diagnostic analysis did not implicate immune cells or pathways ^1^.

Most genetic variants associated with psychiatric risk are in non-coding regions of the genome, likely exerting their effects by altering the activity of regulatory elements ^39^ such as promoters or enhancers; and enhancers can be linearly distant (>10 kilobases) from the genes they regulate ^40^. Some regulatory elements control gene expression in multiple tissues, but others are specific to particular tissues, or particular cell states. For example, some enhancers are active in stimulated but not resting immune cells ^41-43^. The locations and activity status of putative enhancers and promoters in a given tissue can be identified through characteristic epigenetic modifications, such as histone modifications.

Epigenetic mechanisms have long been thought to be important in psychiatry, especially in mediating gene-environment interactions ^44,45^. Epigenetic data from brain tissues have been extensively used to investigate the brain cell types and regions implicated by psychiatric risk variants ^46-48^, by testing whether risk variants tend to be concentrated, or “enriched”, in regions of the genome that are active in a given tissue. However, the enrichment of psychiatric risk variants in immune cell subsets has not been extensively explored. Studies to date have tended to use functional information from whole blood or immune organs, which obscures and dilutes possible effects in the myeloid and lymphoid immune cell subsets comprising these samples. There is some evidence of enrichment of risk variants for bipolar disorder in genes characteristic of neutrophils, T cells and haematopoietic stem cells; and for schizophrenia at genes in T cells and chromatin marks in T and B cells^49^. To our knowledge, no studies have demonstrated enrichment of trans-diagnostic risk, or of cis-risk for MDD or ASD, in any immune cell type ^1,50-52^, or tested if immune cell enrichment is independent of brain tissue enrichment (rather than simply due to coincidental overlap of active genomic regions in brain and immune system cells).

We hypothesized that some genetic risk variants for psychiatric disorders act via their effects on regulatory elements in specific immune cell subsets, thus potentially modulating the response of these cells to infections and other environmental stimuli. We further hypothesized that some of these immunogenetic mechanisms may represent a common pathogenic pathway to multiple psychiatric disorders. To test these hypotheses, we integrated data on common genetic variants associated with trans- and cis-diagnostic risks for psychiatric disorder(s) with data on epigenetically active genomic regions in multiple human cell and tissue types. More formally, we tested the null hypothesis that a given set of risk variants was not co-located with tissue-specific marks of epigenetic activation more frequently than expected by chance in each of multiple tissues (Roadmap/ENCODE ^53,54^), in 19 sorted immune cell subsets (BLUEPRINT ^55^), and in *ex vivo* stimulated naïve and memory CD4^+^ T cells and macrophages (Soskic dataset ^42^). To contextualise our results, we conducted parallel analyses of three “positive control” disorders: Alzheimer’s disease, a brain disorder for which genetic risk has been associated with myeloid immune cells ^56^; rheumatoid arthritis, a canonical adaptive autoimmune disorder; and body mass index (BMI), a common comorbidity which may contribute to observed immune abnormalities in psychiatric disorders ^57^.

## Results

### Trans-diagnostic psychiatric risk is enriched at active chromatin states in T cells

For trans-diagnostic risk of having any one of 8 major psychiatric disorders, we tested for enrichment of genetic risk at active regulatory elements in 88 cells or tissues from the Roadmap consortium, using stratified linkage disequilibrium score regression (s-LDSC). S-LDSC is used to test whether SNP-heritability for a disorder is concentrated or enriched in a genomic annotation^58^. To generate a single binary annotation of active regulatory elements for each tissue, we combined annotations for active promoters and enhancers based on histone marks (see **Methods**). We found that three main tissue classes were significantly enriched for trans-diagnostic risk variants at regulatory elements, following correction for multiple comparisons: multiple adult and fetal brain regions; T cells; and pancreatic islets (**Figure 1A, S1, Table S4**).

**Figure 1.**
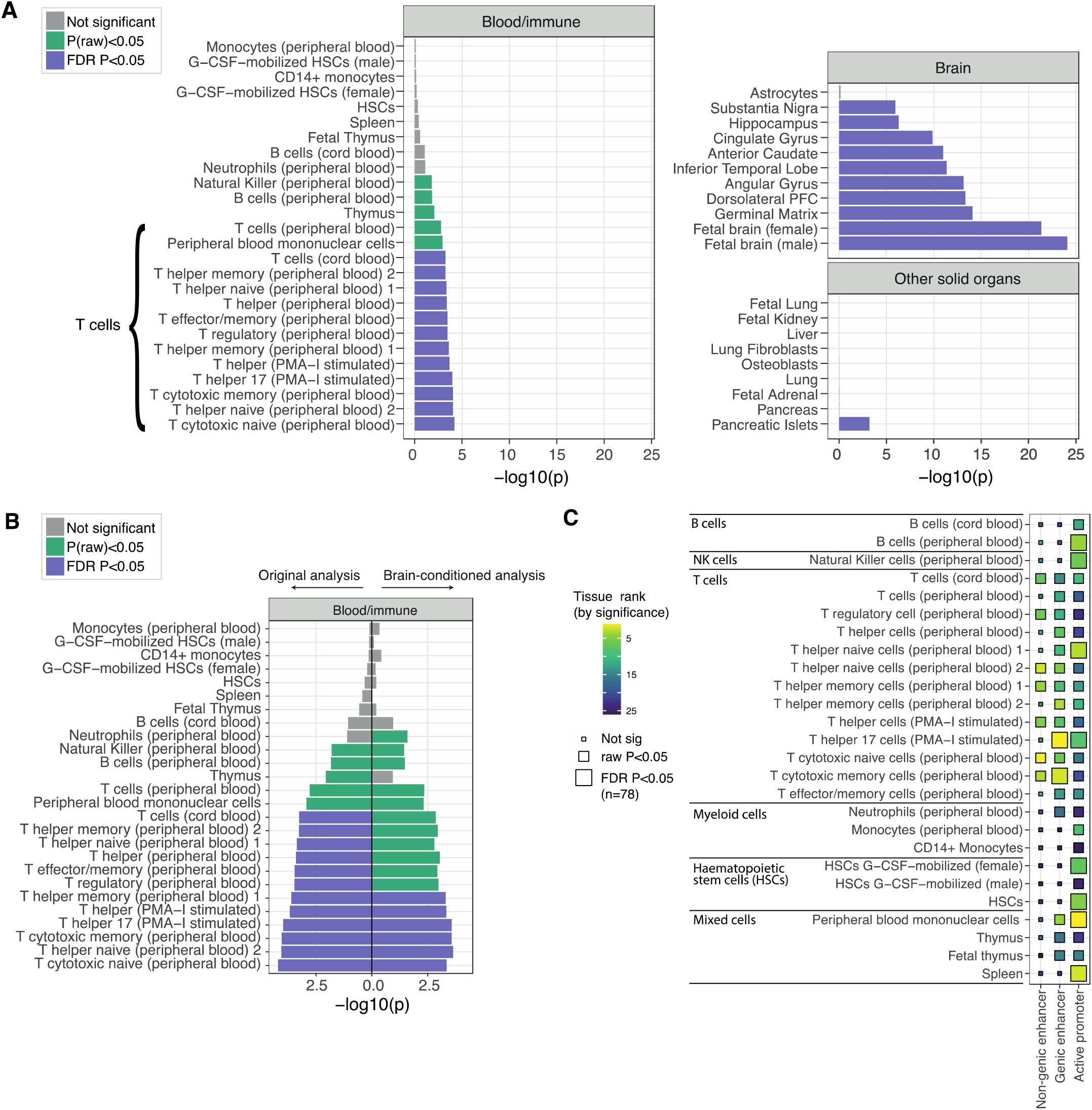
Trans-diagnostic risk enrichment at epigenetically active sites in brain tissue and, independently, in T cells. (A) Enrichment of trans-diagnostic risk at active regulatory elements (active promoters and enhancers) in 88 tissues from the Roadmap epigenomics consortium. *P*-values are shown for the results of stratified linkage disequilibrium score regression (s-LDSC) analysis, taking the union of active elements in a given cell type as the annotation of interest (see **Methods**). The *P*-values from s-LDSC were used to test the null hypotheses that risk variants were not co-located with epigenetically activated sites more frequently than expected by chance, using the Benjamini-Hochberg false discovery rate to correct for multiple tests across N=88 tissues, *P*_FDR_ < 0.05 (purple). Tissues with nominally significant enrichment (*P* < 0.05, green) are also shown for context. Only tissue groups with at least nominally significant results are shown; for non-significant results in other tissue groups (gastrointestinal; skin; fat; muscle; reproductive; vascular) see **Figure S1**. PFC, prefrontal cortex; HSC, hematopoietic stem cells. (B) Brain-conditioned analysis: right-pointing bars show repeat of the analysis in **Figure 1A**, including the active regulatory annotations for all 10 significantly enriched brain regions as additional terms in each of the s-LDSC models for all other cell types. Left-pointing bars show the original results, as in **Figure 1A**. Following conditional analysis, there was no significant decrease in enrichment significance for any immune tissues (*P* > 0.05). Only immune cell results are shown; see **Figure S2B** for conditional results for other tissues. (C) Enrichment of trans-diagnostic risk in immune cell enhancers, genic enhancers and active promoters in Roadmap immune tissues. Tile size indicates s-LDSC significance: large tiles show results significant at *P*_FDR_ < 0.05, using the Benjamini-Hochberg procedure to correct for the 78 annotations tested; mid-sized tiles show nominally significant results (*P* < 0.05) for context. Tile fill indicates the *P*-value rank within each annotation across cell types.

In the central nervous system (CNS), trans-risk variants were most strongly enriched at regulatory elements in fetal brain tissue samples. There was also significant enrichment (*P*^FDR^ < 0.05) at active regulatory elements in brain structures previously reported as abnormal in neuroimaging studies of psychiatric disorders: dorsolateral prefrontal cortex, angular gyrus, inferior temporal lobe, anterior caudate, cingulate gyrus, hippocampus and substantia nigra. In contrast, trans-risk was not enriched at epigenetically active sites in astroglia, consistent with previous analyses demonstrating that genes mapped to trans-risk genetic variants are mainly expressed by neurons^1^ rather than glia or non-neuronal cells in the brain.

In the immune system, trans-risk variants were significantly enriched (*P*_FDR_ < 0.05) at epigenetically active genomic sites in multiple T cell subsets, including cytotoxic, helper and regulatory T cells in adult blood and T cells in cord blood. Conversely, there was no enrichment (*P* > 0.05) of trans-risk in myeloid cells (monocytes, neutrophils).

Many regulatory elements are common to multiple tissues, so we reasoned that this pattern of CNS and immune system enrichment for trans-risk variants could be driven by coincidental overlap of brain and T cell active elements. In this case, the genetic risk would be theoretically expected to have pathogenic effect primarily by its modulation of epigenetically active sites in the brain, with no clearly independent pathogenic role mediated by T cells. We therefore repeated the s-LDSC analysis but included the active annotations for all 10 significantly enriched brain regions as extra terms in the s-LDSC models for every other cell type. In this brain-conditioned analysis, both helper and cytotoxic T cells remained strongly enriched for trans-diagnostic genetic risk (**Figure 1B**), while pancreatic islets did not (**Figure S2B**). For enriched immune tissues in the original analysis (at *P*_FDR_ < 0.05), none showed significantly decreased enrichment following brain-conditioned analysis (two-sample *Z*-test, *P* > 0.05). Conversely, including the annotation for male fetal brain (the brain tissue showing strongest enrichment) as an extra term in s-LDSC models significantly reduced trans-risk enrichment in all other brain regions (*Z*-test *P* < 0.05) except the substantia nigra (*P* = 0.09) and hippocampus (*P* = 0.07), reflecting some overlap of active elements between different brain regions at different developmental phases, and validating our statistical approach (**Figure S2A, Table S3**). We showed the same effect for female fetal brain, the second most strongly enriched brain tissue (**Figure S2A**), excluding a potential effect of sex differences in brain development.

The global active annotation used as a binary marker of epigenetic activation combines trans-risk enrichment at three different classes of regulatory elements: active promoters, genic enhancers (enhancers found in gene bodies), and non-genic enhancers. To identify which classes were most enriched for trans-risk, we tested each class separately and found that the enrichment of trans-risk observed in terms of the global active annotation in T cells was not driven by a single class of regulatory element: there was enrichment of trans-risk at both active promoters (*P*_FDR_ < 0.05) and enhancers (*P*_FDR_ < 0.05 for genic enhancers) (**Figure 1C**).

### Cis-diagnostic risk is enriched at active chromatin states in T cells

Using data from the Roadmap Epigenomics Consortium, we next investigated the enrichment of cis-diagnostic risk variants at epigenetically active sites in brain tissues and immune cells for each of 5 mental health or neurodevelopmental disorders (schizophrenia, bipolar disorder, MDD, autism and ADHD) and each of 3 positive control disorders (Alzheimer’s disease, obesity [BMI], and rheumatoid arthritis).

In the CNS, cis-risks for adult-onset mental health disorders (schizophrenia, bipolar disorder, MDD) were enriched in multiple fetal and adult brain tissues, and cis-risks for child mental health or neurodevelopmental disorders (autism, ADHD) were enriched more selectively in fetal brain tissue. Cis-risk for obesity (BMI) was also enriched for active sites across multiple fetal and adult brain tissues; but cis-risk for Alzheimer’s disease was only (nominally) significantly enriched in hippocampus; and cis-risk for rheumatoid arthritis was not enriched in any brain tissue (**Figure 2A, Table S4**).

**Figure 2.**
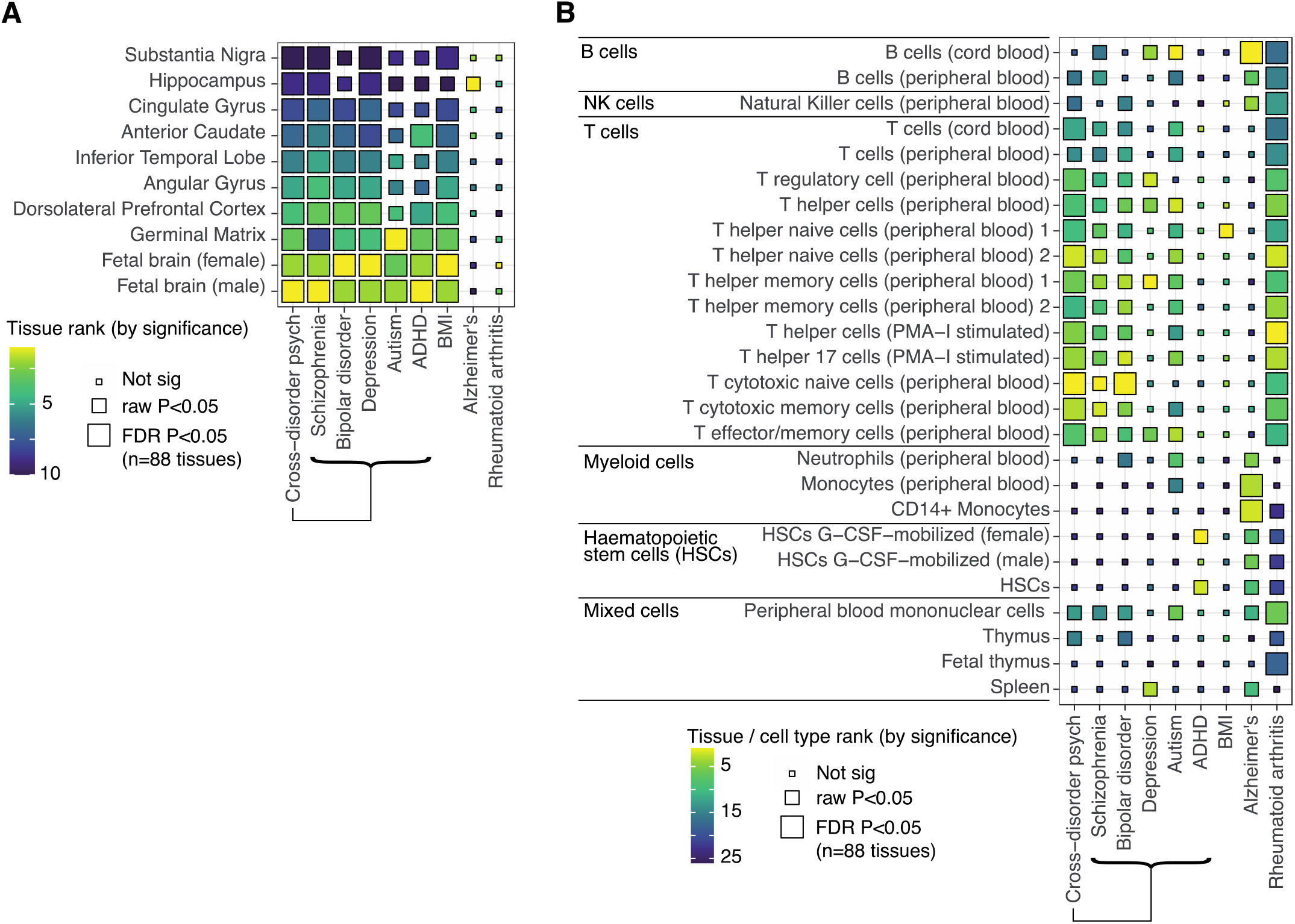
Cis-diagnostic risk enrichment at epigenetically activated sites in adult and fetal brain tissue and immune cells for 8 specific disorders. For each of 5 mental health disorders (schizophrenia, bipolar disorder, major depressive disorder [MDD], autism, and attention deficit-hyperactivity disorder [ADHD]), and for each of 3 positive control disorders (obesity, Alzheimer’s disease and rheumatoid arthritis), enrichment of cis-risk variants at active regulatory elements (active promoters and enhancers) was tested in (A) 10 brain tissue samples (3 fetal) and (B) 26 immune cell classes (3 fetal) ^54^. *P*-values are shown for the results of stratified linkage disequilibrium score regression (s-LDSC) analysis, taking the union of active elements in a given cell type as the annotation of interest. Tile size, from large to small, indicates P-value thresholds from *P*_FDR_ < 0.05 (significant after Benjamini-Hochberg correction for all 88 tissues tested, including those not shown here), through *P* < 0.05 (nominally significant), to *P* ≥ 0.05 (not significant). Tile fill indicates the *P*-value rank within each disorder across all cells/tissues to facilitate comparisons across results from differently-powered genetic association studies.

In the immune system, cis-risks for schizophrenia, bipolar disorder, MDD and autism were enriched at globally activated sites in one or more T cell subsets (mostly at *P* < 0.05; **Figure 2B, Table S4**), with signal driven by both enhancers and promoters (**Figure S3A**). Cis-risk for rheumatoid arthritis was strongly enriched at globally active sites in multiple immune cell subsets; cis-risk for Alzheimer’s disease was significantly enriched in myeloid cells and B cells ^56,59^; and cis-risk for BMI was only enriched in one T cell class at *P* < 0.05 (**Figure 2B**). The statistical significance of these results depends partly on the sample size of the underlying GWAS and the heritability and polygenicity of the disorder (factors influencing power, and captured by the SNP-based heritability *Z*-score)^58^; but also on the strength of functional enrichment of the phenotype in that annotation. For the two most enriched immune and brain annotations (naïve cytotoxic and helper T cells; fetal male and female brain), we tested the correlation between heritability *Z*-score and functional enrichment *Z*-score across the 9 disorders included in this study (**Figure S3B**). We found a strong relationship between heritability *Z*-score and brain enrichment (fetal male brain: Spearman’s ρ = 0.87, *P* = 0.005; fetal female brain: ρ = 0.87, *P* = 0.005), but no correlation between heritability *Z*-score and immune enrichment (cytotoxic T cells: Spearman’s ρ = 0, *P =* 1, helper T cells: ρ = 0.03, *P =* 0.9), suggesting that GWAS power was not driving these results.

### Trans- and cis-risk variants are enriched at active enhancers/promoters in lymphoid cells: BLUEPRINT data

To assess the generalizability of these results in an independent dataset, we tested for enrichment of trans- and cis-risk variants at active enhancer/promoter marks (H3K27ac) in sorted immune cell subsets from the BLUEPRINT consortium ^55^, using the CHEERS algorithm ^42^ to distinguish effects in epigenetically similar cell types (see **Methods**). We replicated key prior findings in the Roadmap data, i.e., trans-risk and cis-risk for schizophrenia and depression were significantly enriched at epigenetically active sites in lymphoid cells; but not myeloid cells (**Figure 3A**). This was not due to problems with the myeloid data, as we detected the expected enrichment of Alzheimer’s Disease risk variants in macrophages **(Figure 3A**). As well as T cell enrichment, we also find enrichment of trans-risk and cis-risk for schizophrenia and (especially) depression in B cells, as well as enrichment of trans-risk and cis-risk for schizophrenia in NK cells. For ADHD and bipolar disorder (less well-powered GWAS studies with fewer independent significant loci available for analysis, see **Table S1**), no cell types were enriched at *P*_FDR_ < 0.05 (**Figure S4A**). Despite both schizophrenia and depression showing strong lymphoid enrichment, the specific histone peaks overlapped by risk variants for these disorders were not generally shared between them (**Figure 3B, S4B**). This indicates that cis-risks for these two disorders were convergently enriched at a cellular level but distinct at the level of specific regulatory elements. It was also notable that cis-risk variants for obesity overlapped with a set of H3K27ac sites that was largely disjoint with the sets of regulatory elements overlapping with cis-risk variants for psychiatric disorders (**Figure 3B**).

**Figure 3.**
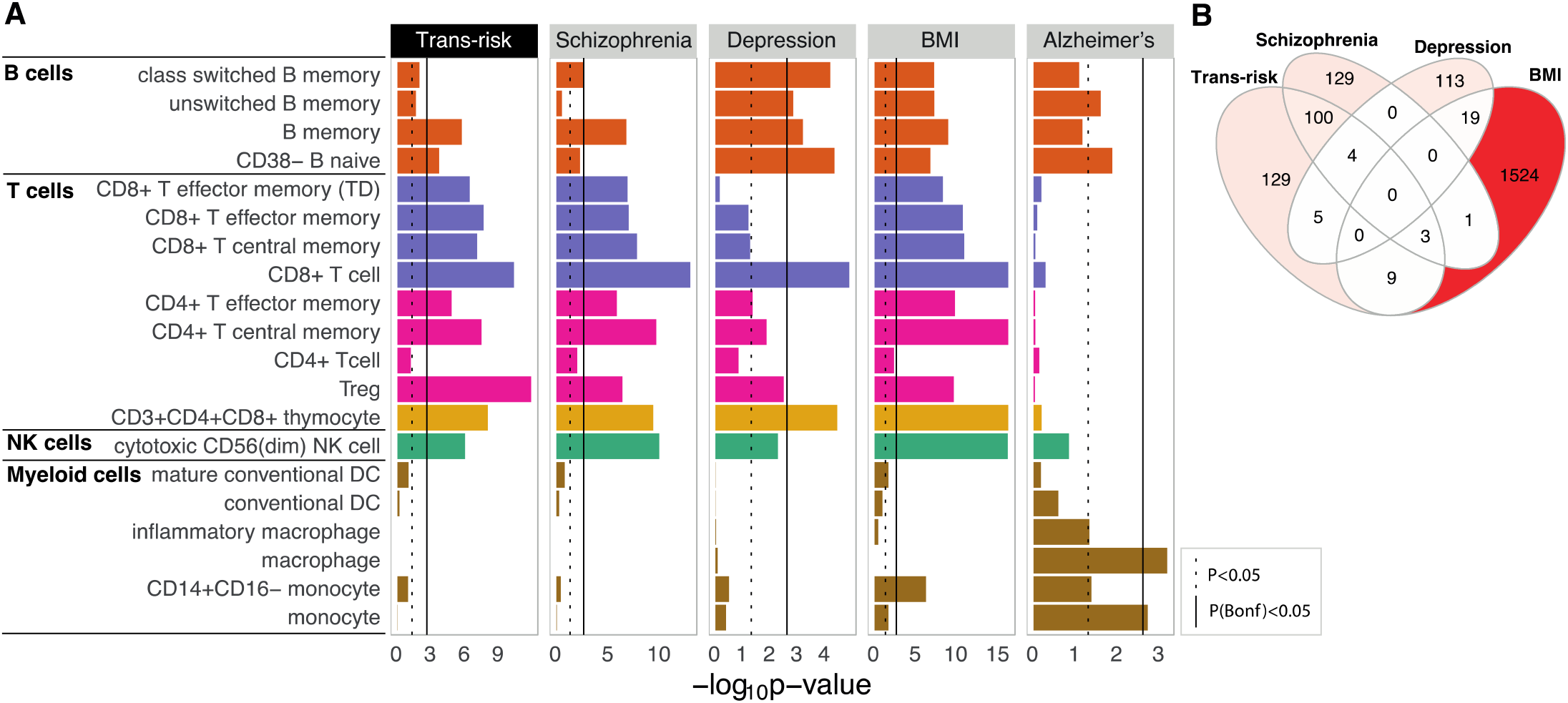
Trans- and cis-diagnostic risk variant enrichment at histone-acetylated marks on adult immune cells in the BLUEPRINT dataset. (a) Bar plots show enrichment of genetic risk for each disorder at active promoters/enhancers (H3K27ac marks) in unstimulated, sorted immune cells. CHEERS was used to detect enrichment of risk loci at cell-type specific H3K27ac peaks (see **Methods**). The dotted black line marks the nominal significance threshold, *P* < 0.05; the solid black line marks the Bonferroni-corrected significance threshold, *P*_FDR_ < 0.05. Note differing x-axis scales. See **Figure S4A** for ADHD and bipolar disorder results (non-significant after FDR control). (b) Venn diagram shows counts of variant-peak overlaps shared between disorders and unique to each disorder (each peak is only counted once even if overlapping multiple variants). For an upset plot of peak overlaps across all disorders, see **Figure S4B**.

### Trans- and cis-diagnostic risk variants are enriched at histone-acetylated sites in stimulated T cells: Soskic immune stimulation dataset

Given that risk of mental health disorders is affected by both genetic variation and environmental factors, we reasoned that trans- and cis-risk variants could be most significantly enriched at sites that were epigenetically activated in immune cells stimulated by cytokines (mimicking environmental insults) towards different activated cell fates. To investigate this hypothesis, and to assess the robustness of our principal findings in a third independent dataset, we tested whether trans- and cis-risks were enriched at regulatory elements (H3K27ac marks) active during immune cell activation, using a dataset of human naïve and memory CD4^+^ T (helper) cells and macrophages stimulated *ex vivo* in the presence of 13 cytokine combinations. The chromatin activity was assessed at early and late timepoints after exposure to cytokine stimulations (16h and 5 days for T cells and 6h and 24h for macrophages), as well as in unstimulated cells ^42^. Both trans-diagnostic risk variants, and cis-risk variants for MDD were most significantly enriched in memory T helper cells at day 5 following T cell stimulation with anti-CD3/anti-CD28 beads that mimic activation occurring with T cell receptor-crosslinking; trans-risk variants and cis-risk variants for schizophrenia were also significantly enriched in memory T helper cells at 16 h and in naïve T helper cells at day 5 only (**Figure 4A**). The loci of histone acetylation that overlapped with cis-risk variants for MDD in late-activated memory T cells were almost completely disjoint with the set of epigenetic marks that overlapped with cis-risk variants for schizophrenia in late-activated memory T cells (**Figure S6A**), again demonstrating convergence of cis-risk enrichment at the immune cell subset level, but divergence at the molecular level of specific regulatory elements. Enrichment was generally much greater for stimulated than unstimulated T cells, with smaller differences in the magnitude of enrichment between different cytokine stimulation conditions (**Figure 4A**). For 9 of the 10 cytokine conditions (all except Th17-cytokine polarizing condition), trans-risk enrichment was significantly greater (*Z*-test *P* < 0.05) in stimulated compared to unstimulated late-activated memory T cells.

**Figure 4.**
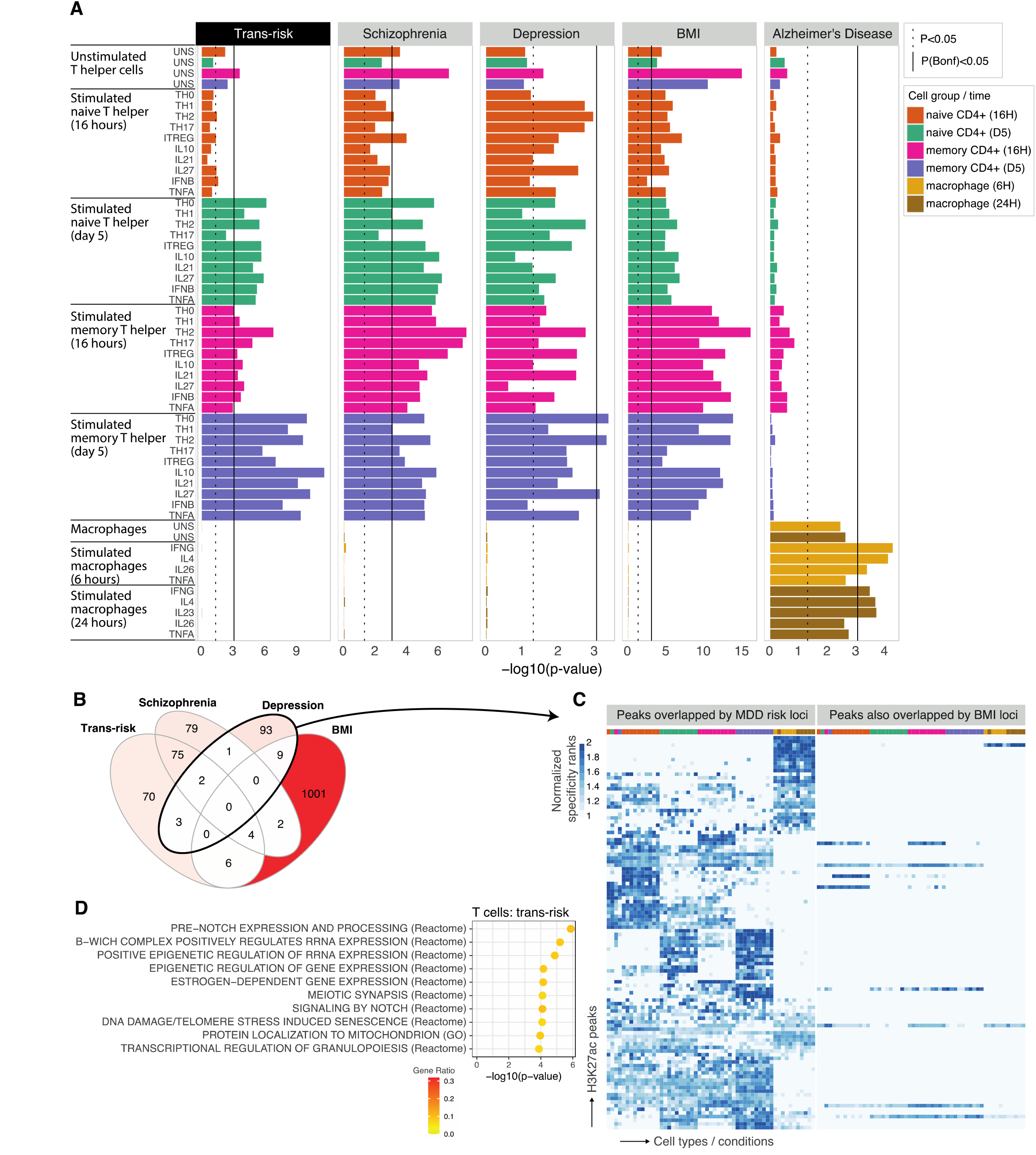
Trans- and cis-risk variant enrichment at histone-acetylated marks on experimentally stimulated immune cells in the Soskic immune stimulation dataset. (A) Bar plots show enrichment of genetic risk for each condition at active promoters/enhancers (H3K27ac marks) in sorted and unstimulated or *ex vivo* stimulated immune cell classes: macrophages, naïve CD4^+^ (helper) T cells and memory CD4^+^ T cells, assayed at both early and late timepoints after stimulation with one of several different cytokine cocktails promoting differentiation to different T cell states (as shown in row labels). CHEERS was used to detect enrichment of risk loci at cell-type specific H3K27ac peaks (see **Methods**). The dotted black line marks the nominal significance threshold, *P* < 0.05; the solid black line marks the Bonferroni-corrected significance threshold, *P*_Bonferroni_ < 0.05. Note differing x-axis scales. Results for other disorders are shown in **Figure S5**. (B) Venn diagrams show counts of variant-peak overlaps shared between disorders and unique to each disorder. For an upset plot of peak overlaps across all disorders, see **Figure S6B**. (C) All Soskic immune stimulation dataset peaks overlapped by risk variants for major depressive disorder. Each row corresponds to an H3K27ac peak overlapping a risk variant for MDD; each column corresponds to a different cytokine-induced cell state, ordered and colored as in **Figure 4A** (see legend). The blue fill shade represents how specific each peak is to each cell state (specificity rank of each peak normalized to the mean specificity rank of all peaks). Only 9 of the 108 MDD-associated H3K27ac immune peaks also overlap BMI risk variants. (d) For peaks which were both highly specific to T cells (including both unstimulated and stimulated cells) and overlapped by trans-risk variants, nearest genes were identified and tested for enrichment for curated biological pathways (GO and Reactome) using a hypergeometric test. Only the 10 most significant pathways are shown (all *P*_FDR_ < 0.05). Fill colour indicates gene ratio (number of test genes in the pathway / total number of test genes). See **Figure S7** for results for cis-diagnostic risks.

As in the two prior independent datasets, there was no enrichment for trans-or cis-risk of psychiatric disorders at epigenetically activated sites in myeloid cells, either stimulated or unstimulated, with the exception of enrichment of bipolar disorder risk in IL26-stimulated macrophages (**Figure S5**). Cis-risk variants for obesity were enriched in unstimulated and stimulated T cell states (**Figure 4**), but only 9 of the 108 depression-associated H3K27ac peaks also overlapped with BMI risk variants (**Figure 4C, S6B**), indicating that cis-risks for obesity and depression were enriched at distinct regulatory elements in the same cell subsets.

For disorders showing enrichment in T cells, we performed pathway analysis (overrepresentation analysis) for those genes overlapping or nearest to the T-cell specific histone acetylation peaks overlapped by risk variants. Trans-risk (**Figure 4D**) and cis-risk for schizophrenia (**Figure S7**) showed enrichment of pathways including epigenetic regulation, pre-notch processing and estrogen-dependent gene expression in T cells. In contrast, rheumatoid arthritis showed enrichment of lymphoid cell differentiation, activation, and response to antigenic stimulus (**Figure S7**).

## Discussion

We examined the enrichment of genetic risk variants for psychiatric disorders at epigenetically activated regulatory sites across multiple tissues. Trans-diagnostic risk variants, commonly associated with multiple mental health and neurodevelopmental disorders, were significantly enriched at active regulatory sites in several adult and fetal brain tissue samples. Strikingly, we also found that trans-diagnostic risk variants were significantly enriched at an independent set of regulatory elements in peripheral blood lymphoid cells (but were not enriched in myeloid cells). Our key results – enrichment of trans-risk in T cells and lack of enrichment in myeloid cells – were statistically robust to multiple comparisons and replicated in three independent datasets. Other lymphoid cells (for which fewer datasets were available) are likely also implicated in pathogenesis, as we also found enrichment of trans-risk in B cells and NK cells.

Further investigation of cis-diagnostic risk variants, specifically associated with one of 5 mental health or neurodevelopmental disorders, confirmed significant enrichment of genetic risks for schizophrenia and major depressive disorder at active promoters and enhancers in both brain tissue and peripheral lymphoid cells (but not myeloid cells). Epigenetically activated sites in T cells, especially cytokine-stimulated CD4^+^ T cells, were most consistently and significantly enriched for sequence variants associated with schizophrenia or MDD; however, the active regulatory elements overlapped by these cis-diagnostic variants were specific to each disorder. This suggests convergence of risk for schizophrenia and depression at a cellular level in the immune system, i.e. activated T cells, and raises questions about how the involvement of different specific regulatory elements in these two disorders might relate to the different phenotypic presentations of schizophrenia and depression. We also found strong enrichment of risk for depression in both naïve and memory B cells. To our knowledge, this is the first demonstration of enrichment of genetic risk for MDD at epigenetically active sites in lymphoid cells (or indeed any immune cell type).

The cis-diagnostic enrichment results for schizophrenia and MDD were statistically robust to multiple comparisons and in clear contrast to the comparable results for 3 positive control disorders. Cis-risks for Alzheimer’s disease were significantly enriched at epigenetically activated sites in myeloid cells (but not lymphoid cells); cis-risks for rheumatoid arthritis were enriched at active sites in myeloid and lymphoid cells (but not brain tissue); and cis-risks for obesity (BMI) were enriched at active sites in brain tissue and (in some analyses) in immune cells, but with effects on regulatory elements distinct from those implicated by psychiatric disorders.

On this basis, we propose that genetic variants associated with increased risk for psychiatric disorders are likely to interact with epigenetic activation of specific and distinct regulatory elements in both the central nervous system and the adaptive immune system. This pathogenic model immediately raises three key questions. What environmental exposures cause epigenetic modification at risk-enriched sites in T cells? How could atypical T cell phenotypes, arising from this gene-by-environment interaction, cause changes in the CNS that are ultimately manifest as mental health or neurodevelopmental disorders? What is the antigen presenting cell which activates atypical CD4^+^ T cells (our data suggest this may be B cells)?

Infection is the most likely environmental stimulus to induce epigenetic activation in the immune system. And there is abundant epidemiological evidence that fetal and post-natal infections increase the risk for multiple psychiatric disorders ^26,27,30,31^. However, the immune mechanisms by which early-life infection predisposes to later psychiatric symptoms are not known. We know that fetal or childhood infections can cause long-term changes in adaptive immune cell phenotypes, including T cell memory of antigens and B cell production of antibodies, that are crucial to development of adult immunity. Thus, it is conceivable that the epigenetically activated sites enriched for trans- and cis-risks in T cells and memory B cells in these data were “marked” by exposure to infection or inflammation; and that genetic risk variants modulate the infection-induced activation of regulatory elements, leading to atypical T or B cell phenotypes following infection in people at genetic risk of psychiatric disorder. There is already some epidemiological evidence for gene-by-environment interactions between infection and risk variants for schizophrenia ^60-62^ and MDD ^63^. Many aspects of our data are compatible with this concept. For example, our finding that trans- and cis-diagnostic risk variants are enriched at sites epigenetically activated by delayed T cell responses to a wide range of pro-inflammatory cytokine stimuli seems consistent with the epidemiological finding that increased risk of multiple psychiatric disorders is found following a wide range of different infections ^12,28,29,64,65^.

Atypical T cell phenotypes could conceivably have effects on the brain by at least two broad routes: via stimulus-driven T cell activation and via developmental pathways (**Figure 5**). By this account, the (epi)genetic root cause of mental health-related inflammation is located in the adaptive immune system, rather than the innate immune system. Atypical T cells are supposed to impact on neuronal function via the well-documented adverse effects of soluble inflammatory mediators ^66^; via contact-dependent mechanisms; or via their effects on other immune or non-immune cells which in turn affect neurons. In contrast with autoimmune diseases, which tend to show greatest enrichment in early T cell activation states ^42^, the strongest enrichment for psychiatric risk variants was in T cells, especially late-activated memory CD4^+^ T cells, and memory B cells. This may reflect abnormalities in the resolution (rather than onset) of immune responses, potentially leading to chronic, low-grade peripheral inflammation seen in many psychiatric disorders.

**Figure 5.**
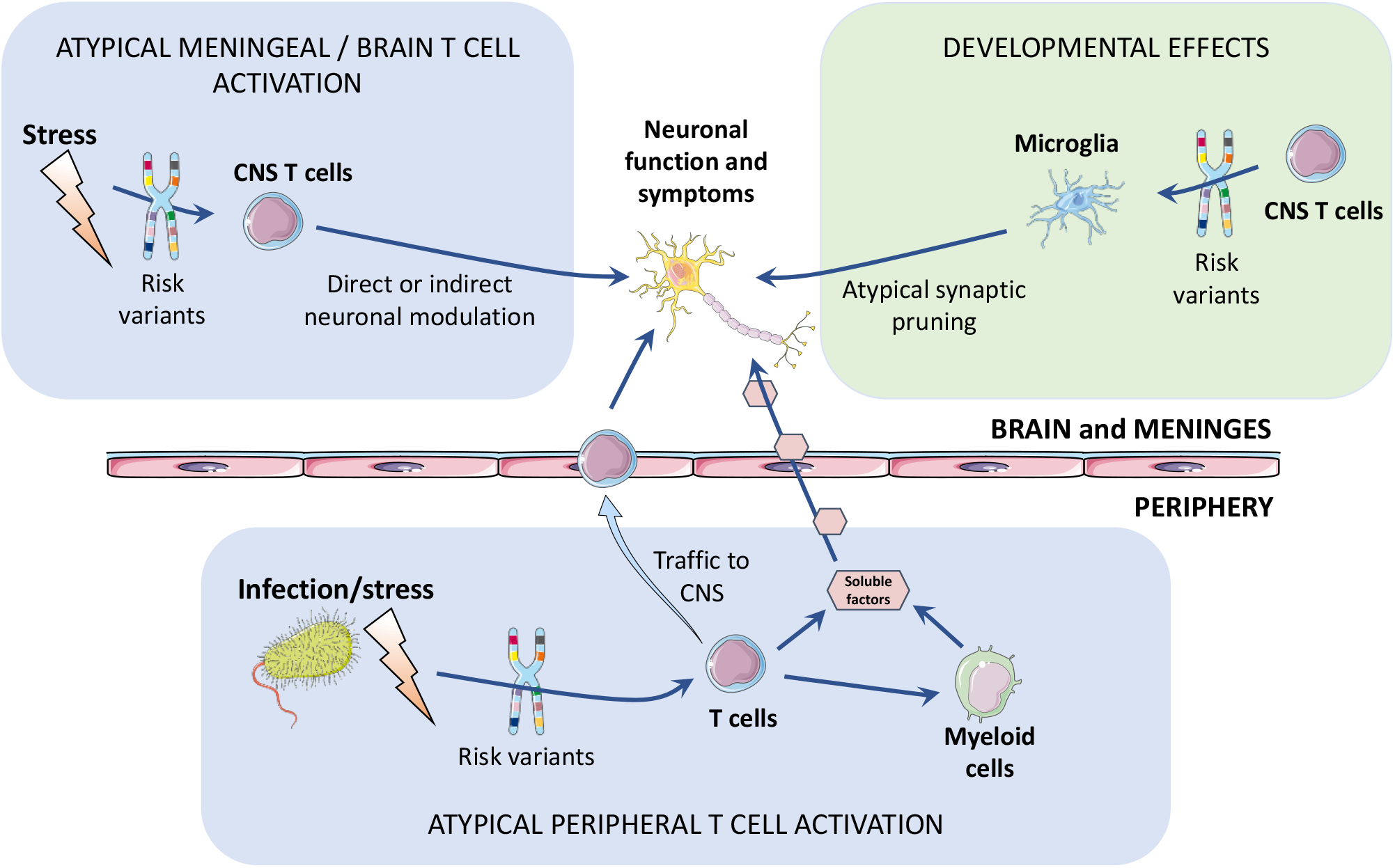
Schematic of potential pathogenic pathways by which genetic risk variants enriched at epigenetically active sites in T cells could lead to neuronal changes and ultimately psychiatric disorders. Atypical stimulus-driven activation pathway (light blue boxes): Infection or other stressors can induce epigenetic activation of regulatory elements in T cells that are enriched for trans-or cis-diagnostic risk variants, proximally causing atypical T cell phenotypes, and distally causing increased inflammatory activation of innate immune (myeloid) cells in the periphery and CNS. Atypical activation of T cells resident in the meninges or brain, or trafficking into the meninges and brain from the periphery, could directly have adverse effects on neuronal function. Developmental pathway (light green box): T cells typically control microglial pruning of neuronal synapses as part of normative brain developmental programs in childhood and adolescence. Atypical T cells, potentially induced by infection or stress in genetically-at-risk individuals, could promote atypical microglial pruning of synapses, contributing to the formation of disconnected networks or circuits in the adult brain.

The developmental pathway follows growing evidence that T cells – even in the absence of infection – have an important role in controlling the activity of microglia, the brain’s most numerous resident myeloid cells, in phagocytosing synaptic terminals and neurites as part of normative neurodevelopmental programs of synaptic pruning that are ongoing from early childhood throughout adolescence. Thus atypical T cells in the meninges or brain could lead to atypical synaptic pruning ^67,68^, mediated by innate immune cells (microglia), with long-term consequences for maturation of large-scale brain networks of inter-connected neurons. There is evidence already from neuroimaging studies that schizophrenia and other psychiatric disorders are associated with disrupted development of connectivity in cortico-subcortical circuits ^69^. It will be important in future studies to investigate more directly the hypothesis that genetic risks for psychiatric disorders can result in development of dysconnected brain networks via immune as well as neuronal mechanisms.

Perhaps surprisingly, given the prior focus on innate immune abnormalities associated with psychiatric disorders, epigenetically active sites in myeloid cells were not significantly enriched for trans-risk variants or for cis-risk variants for schizophrenia or MDD. What does this mean for the pathogenic role of myeloid cells in these disorders? It is possible that genetic risk variants are indeed enriched at epigenetically active sites in myeloid cells, but only under stimulation conditions that were not represented in the three datasets we analysed. Alternatively, it could be that activation of myeloid cells and associated release of CRP and pro-inflammatory cytokines, although commonly measured as immune biomarkers in previous studies of psychiatric disorders, are downstream consequences of an atypical helper or regulatory T cell response to infection or stress. This would be consistent with the well-known role of T cells in coordinating and modulating innate immune function ^70-72^, including via inflammasome inhibition ^73^. Thus atypical T cell phenotypes could promote or sustain low-grade inflammatory states of the innate immune system in the periphery, in the meninges, and in the brain, that have been robustly associated with MDD, schizophrenia and other psychiatric disorders ^74-76^. A third possibility is that inflammatory responses of myeloid cells have a causal role in pathogenesis of psychiatric disorders but this is entirely driven by environmental factors and independent of inherited genetic risks.

The statistical significance of results for risk variant enrichment at epigenetically active regions reflects in part the sample size of the GWAS’s used and the heritability and polygenicity of the disorders. However, in contrast to our results for brain enrichment, we did not find any correlation between GWAS statistical power and immune enrichment. This suggests that while GWAS power can explain some differences in functional enrichment (as in brain), differences in immune cell enrichment may be more indicative of between-disorder differences in how strongly immune mechanisms contribute to pathogenesis in general, or how frequently immune mechanisms are implicated in individual patients clinically diagnosed with a specific disorder.

We focused here on European ancestry genetic results, as the currently available datasets are from European participants, but the immunogenetics of psychiatric risk should be examined in other ancestries. In addition, the epigenetic datasets used here are predominantly adult: given the role of developmental insults in psychiatric risk, it will be important to investigate genetic enrichment in immune cells sampled at different developmental stages, including adolescence.

Likewise the immune cell states considered in this analysis are the canonical states associated with infection and autoimmunity. It will also be important to explore whether genetic risk variants modulate immune cell phenotypes induced by exposure to non-infectious environmental stimuli e.g. stress, especially given that childhood adversity and other social stressors are known to profoundly increase risk for multiple psychiatric disorders.

In psychiatry, T cell phenotypes have been most investigated in schizophrenia ^77^, with some evidence of decreased proliferative responses to stimulation ^78^. Our findings motivate further investigation of T cell and B cell phenotypes across multiple psychiatric conditions, with a focus on how trans-risk affects activated T cells (e.g. stimulated ex-vivo). Functional genomic analysis of T cell subsets from patient cohorts will be particularly important to directly test for disease-associated alterations in DNA accessibility, histone modifications, enhancer-promoter interactions and gene expression. We hypothesize that these alterations will be found at those T cell peaks identified in our analysis as overlapping risk SNPs. However, there may be broader epigenetic consequences of risk variants, especially given that pathway analysis implicated epigenetic regulation processes, which may occur at sites distant from the risk variants.

In conclusion, genetic risk variants for psychiatric disorders were significantly enriched at epigenetically active enhancers/promoters in adaptive immune cells, especially stimulated T cells. This suggests a mechanistic role for T cells in mediating the interaction between environmental exposures and genetic risk variants in the pathogenesis of multiple psychiatric disorders.

### Data sharing statement

All datasets and code used for this analysis are publicly available (see **Table S2**).

## Data Availability

All datasets and code used for this analysis are publicly (see Supplementary Table 2 for full details)
Github repository: https://github.com/maryellenlynall/psychimmgen2021
Processed datasets deposited in Zenodo: https://doi.org/10.5281/zenodo.5153661

https://doi.org/10.5281/zenodo.5153661

## Acknowledgements

M.E.L is supported by the Medical Research Council. This work was additionally supported by the NIHR Cambridge Biomedical Research Centre. E.T.B. is supported by an NIHR Senior Investigator award. B.S. is funded by the Open Targets grant OTAR040. G.T. is supported by the Open Targets and the Wellcome Trust (grant WT206194). DFL is supported by a US Veterans Affairs Career Development Award. The authors would also like to acknowledge the crucial input of the Sanger Institute Cellular Genetics Informatics team.

## Methods

### Trans- and cis-diagnostic genetic risk variants for psychiatric disorders

The primary GWAS datasets used for the identification of trans- and cis-risk genes are listed in **Table S1**. We used summary statistics from a meta-analysis of trans-diagnostic risk across 8 mental health or neurodevelopmental disorders ^1^: ASD, bipolar disorder, MDD, obsessive-compulsive disorder, schizophrenia, anorexia nervosa, ADHD, and Tourette syndrome. For analysis of cis-risk, i.e. risk of a specific psychiatric disorder, we separately tested 5 large primary genome-wide association studies (GWAS) of MDD^79^, bipolar disorder ^52^, schizophrenia^80^, autism^81^, and ADHD ^82^. For comparative purposes, we analysed GWAS results for BMI ^83^, Alzheimer’s disease ^84^, and rheumatoid arthritis ^85^. For all disorders except MDD, we selected the largest publicly available, predominantly-European GWAS dataset; for MDD, we used a larger recent European GWAS ^79^.

### Testing for enrichment of genome-wide genetic risk at regulatory elements (ROADMAP data)

Stratified linkage disequilibrium score regression (s-LDSC) can be used to test whether genetic risk is concentrated or enriched in a genomic annotation, e.g. a set of active regulatory elements in a specific cell type ^58^. S-LDSC hinges on the fact that the disease association statistic for a given genetic variant depends on whether that variant is linked to the disease, but also whether variants in linkage disequilibrium (LD) with that variant are linked to the disease. By testing whether variants in LD with the annotation of interest tend to have higher association scores than variants elsewhere, we can calculate an enrichment score capturing the tendency of SNP-based heritability for that disease to be co-located with that annotation ^58^. We used this method to test for enrichment of psychiatric risk variants at active regulatory elements in 88 cell or tissue types.

For a given tissue, CHiP-seq data assaying multiple histone marks can be integrated to segment the genome into annotations representing different functional epigenetic states, e.g., enhancers, promoters, repressed regions ^54^. The IDEAS algorithm ^86^ leverages shared features across cell types to improve this segmentation. Lacking a strong prior hypothesis about which particular regulatory elements in immune cells would be implicated by psychiatric risk, we generated a simple functional annotation of active states for each tissue in the Roadmap Epigenomics Dataset, which includes samples from all major organ systems including brain, heart, muscle, gut, adipose, skin, reproductive and immune tissues ^54^. Data for a given tissue or cell type sometimes come from multiple donors – as is the case for most of the brain and immune samples – and sometimes from single donors (see https://egg2.wustl.edu/roadmap/web_portal/meta.html for metadata). Immune cell subsets were magnetically sorted from live donor blood samples; brain tissues were homogenized post-mortem samples. For each Roadmap tissue/cell type, we generated a whole genome binary annotation of active regulatory elements (**Figures 1A,1B**) from IDEAS annotations based on 5 epigenetic histone marks (H3K4me3, H3K4me1, H3K36me3, H3K27me3 and H3K9me3). We combined the 6 IDEAS annotations representing active promoters and enhancers to generate a single binary annotation of active regulatory elements for each tissue. More exactly, we merged the IDEAS annotations for active transcription start sites (10_TssA); regions flanking active TSS (8_TssAFlnk); weak TSS (14_TssWk); enhancers (4_Enh); genic enhancers (6_EnhG); and genic enhancers associated with transcription (17_EnhGA), following a previous definition of active states ^87^. We generated partitioned linkage disequilibrium (LD) scores for each tissue as recommended, using HapMap3 SNPs ^58^.

We then used s-LDSC to test the enrichment of psychiatric risk variants in each cell type, using a separate model for each cell type, as is standard. Summary statistics were preprocessed using the LDSC recommended script munge_sumstats.py and we performed s-LDSC for each tissue in the Roadmap dataset, using recommended settings, excluding the MHC regions, and including standard non-cell type specific annotations (baseline v1.2) (see https://storage.googleapis.com/broad-alkesgroup-public/LDSCORE/readme_baseline_versions).

To further dissect the s-LDSC results for the active annotations, we also performed s-LDSC for the 3 types of genomic element comprising the active annotation: promoters, enhancers, and genic enhancers. We generated partitioned LD scores for the promoters (10_TssA, 8_TssAFlnk and 14_TssWk), enhancers (4_Enh) and genic enhancers (6_EnhG and 17_EnhGA) (**Figure 1C, Figure S3A**) then performed s-LDSC using default settings for each of these annotations in the Roadmap immune tissues. The *P*-values from s-LDSC indicate the significance of the coefficient for the cell type specific annotations. *P*-values were corrected for multiple comparisons across tissues using Benjamini-Hochberg false discovery rate. We coloured heatmaps by *P*-value rank to aid comparison across disorders or annotations which are differently powered.

To account for the possible confounding effect of shared regulatory elements between brain and immune tissues, we also performed brain-conditioned enrichment analyses: for each tissue’s s-LDSC model, we added terms for the active regulatory annotations for possibly confounding brain regions. SNP heritability *Z*-scores (heritability / standard error) and s-LDSC *Z*-scores (enrichment coefficient / standard error) were estimated using LDSC. To compare the results of the original and brain-conditional analyses, we used a one-sided two-sample *Z*-test as follows, where *β*_1_ is the coefficient for the annotation in the original analysis and *β*_2_ is the coefficient in the brain-conditional analysis. SE is the standard error of the coefficient for the original (*SE*_1_) or conditional (*SE*_2_) analysis. *Z*-scores were converted to *P*-values.

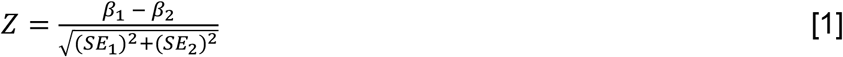

### Testing for enrichment of genetic risk variants in cell-type specific active promoters/enhancers

To compare enrichment of genetic risk at regulatory marks in different immune cell subsets, and immune cells stimulated under different conditions, we used the CHEERS algorithm ^42^. CHEERS quantifies the overlap of lead (independently significant) genetic risk variants with cell-specific epigenetic peaks. Crucially, CHEERS facilitates the comparison of similar cell types or conditions, which tend to have similar epigenetic profiles, by calculating peak specificity scores, indicating how specific a peak is to that cell type relative to other cell types, then quantifying cell-type enrichment as the specificity-weighted sum of overlaps of disease risk variants with these peaks. While s-LDSC leverages genome wide-information, CHEERS focuses on risk loci which meet genome-wide significance. In brief, CHEERS identifies peaks which overlap lead variants or variants in strong LD (r^2^<0.8) with lead variants, then calculates the mean cell type specificity score (in that cell type) of those peaks, which captures the degree of enrichment of that cell type for a given disorder. One-sided *P*-values were reported from a discrete uniform distribution (reflecting the ranking of specificity scores within each cell type) and corrected for multiple comparisons across tissues using a Bonferroni correction. To identify lead disease risk loci, all summary statistics were processed consistently: liftover to hg38, harmonization, removal of MHC region, and distance-based clumping (see below for more detail). We applied CHEERS using two human H3K27ac ChIP-seq datasets: (i) BLUEPRINT consortium data from 19 sorted unstimulated immune cells subsets (see **Figure 3A**) ^55^ and (ii) the Soskic immune stimulation data from sorted and *ex vivo* stimulated immune cells ^42^. H3K27ac marks active (rather than inactive or poised) enhancer and promoter regions ^53,88^. In the Soskic immune stimulation experiment, macrophages, naïve CD4^+^ T cells and memory CD4^+^ T cells were stimulated using different cytokine cocktails associated with autoimmunity or known to promote different cell fates (see **Figure 4A**). In addition, generic T cell receptor and CD28 co-stimulation signals were provided in all stimulated T cell conditions using beads coated with anti-CD3 and anti-CD28 antibodies. H3K27ac data were processed as described previously ^42^ to obtain cell-type specificity scores for H3K27ac peaks in each cell type or state. Here, we ran CHEERS using r^2^ linkage disequilibrium values taken from unrelated European individuals from the 1000 genomes dataset^89^, calculated using PLINK^90^.

To compare the enrichment between stimulated and unstimulated cell subsets, we used a one-sided, two-sample Z-test as follows, where *x*_1_ is the mean specificity rank for the stimulated cell subset and *x*_2_ is the mean specificity rank for the corresponding unstimulated cell subset. SE is the standard error of the mean and depends on the number of SNPs overlapping peaks. For a given disorder, SE is the same across different annotations, as peaks are called across the dataset as a whole. *Z*-scores were converted to *P*-values.

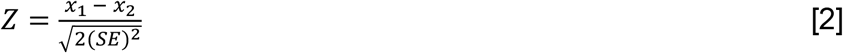

### Identification of independent risk loci

To identify independently significant loci for each disorder, we reprocessed all summary statistics consistently. Given the lack of well-matched linkage disequilibrium data for the populations underlying these studies, we aimed to conservatively identify independent lead variants without using LD information or conditional analysis within loci. We first lifted over the summary statistics (autosomal chromosomes only) and harmonized variants to the reference strand using the EBI summary statistics snakemake pipeline (https://github.com/EBISPOT/gwas-sumstats-harmoniser). Alleles with a minor allele count <10 were filtered out; where minor allele counts were not available, these were imputed from GnomAD v2.1.1 ^91^ European frequencies lifted over to GRCh38. We then filtered all summary statistics to those variants also present at minor allele frequency > 0.01 in 1000 genomes phase 3 (unrelated European participants) called against GRCh38 ^89^. To find independently significant lead loci, we used the Open Targets genetics finemapping pipeline (https://github.com/opentargets/genetics-finemapping) to filter summary statistics to variants with *P* < 5 × 10^−8^ (excluding MHC region chr6:28510120-33480577) and performed distance-based clumping of significant variants with a clumping distance of ±500kb. The number of lead SNPs identified for each disorder is shown in **Table S1**. ASD was excluded from downstream CHEERS analysis as only two significant loci were detected.

### Over-representation analysis

Following CHEERS analysis, to test which biological pathways were implicated in T cells, we identified those T cell-specific peaks overlapped by disease risk variants, selected the genes overlapping or nearest to those peaks, then performed pathway analysis on those genes. More specifically, we selected (for a given disorder) the union of peaks highly specific (CHEERS specificity rank >0.9) to any T cell subset in the Soskic immune stimulation dataset which were also overlapped by risk variants for that disorder. For each peak, we used the ChIPseeker seq2gene function ^92^ to identify those genes overlapping the peak, or with a promoter overlapping the peak, or (if no promoter overlapped the peak) the nearest gene (a maximum of 100 kilobases away). The selected genes were tested for enrichment of GO biological processes and Reactome pathways using a hypergeometric test via the clusterProfiler enricher function, with Benjamini-Hochberg correction for multiple testing ^93^.

