## Supplementary material for "Genetic variants associated with cross-disorder and disorder-specific risk for psychiatric disorders are enriched at epigenetically active sites in peripheral lymphoid cells"

Mary-Ellen Lynall[1,2,3,4], Blagoje Soskic[5,6], James Hayhurst[6], Jeremy Schwartzentruber[5], Daniel F. Levey[7,8], Gita A. Pathak[7,8], Renato Polimanti[7,8], Joel Gelernter[7,8,9], Murray B. Stein[10,11], Gosia Trynka[5,6], Menna R. Clatworthy[3,4], Ed Bullmore[1,2]

[1]University of Cambridge, Department of Psychiatry, Herchel Smith Building of Brain & Mind Sciences, Cambridge Biomedical Campus, Cambridge CB2 0SZ, UK

[2]Cambridgeshire & Peterborough NHS Foundation Trust, Cambridge, UK

[3]Molecular Immunity Unit, University of Cambridge Department of Medicine, Cambridge, UK

[4]Cellular Genetics, Wellcome Sanger Institute, UK

[5]Wellcome Sanger Institute, Wellcome Genome Campus, Cambridge, UK

[6]Open Targets, Wellcome Genome Campus, Hinxton, UK

[7]VA Connecticut Healthcare System, West Haven, CT, USA

[8]Yale Univ. School of Medicine, Dept Psychiatry, New Haven, CT, USA

[9]Yale Univ. School of Medicine, Depts. Of Genetics and Neuroscience, New Haven, CT, USA

[10]VA San Diego Healthcare System, San Diego, CA, USA

[11]Department of Psychiatry and School of Public Health, University of California San Diego, La Jolla, CA, USA

### Contents


- Table S1: Genome-wide association study details
- Table S2: Code and data availability
- Table S3: Statistical comparison of original vs. conditional stratified linkage disequilibrium score regression (s-LDSC models) by *Z*-test
- Table S4: s-LDSC results for active regulatory annotations for 88 ROADMAP tissues for all disorders (statistical tables; separate supplementary excel file)
- Figure S1: Enrichment of trans-diagnostic risk at active regulatory elements (active promoters and enhancers) in 88 tissues from the Roadmap epigenomics consortium
- Figure S2: Cross-disorder psychiatric risk is enriched in brain and lymphoid immune cell active genomic elements
- Figure S3: Enrichment of genetic risk for multiple psychiatric disorders in immune tissues
- Figure S4: Psychiatric genetic risk enrichment at active lymphoid enhancers/promoters (BLUEPRINT dataset)
- Figure S5: Psychiatric genetic risk enrichment at activation-dependent T cell enhancers/promoters (Soskic immune stimulation dataset)
- Figure S6: Soskic stimulated immune cell dataset: overlap of H3K27ac peaks implicated by different disorders
- Figure S7: Soskic stimulated immune cell dataset: pathway enrichment for genes nearest to peaks both specific to T cells and overlapped by risk variants

### Table S1

| **Study** | **Number cases** | **Number controls** | **Number of genome-wide independently significant loci** | **Download link** |
|---|---|---|---|---|
| Cross-disorder psychiatric risk [1] | 162,151 | 276,846 | 115 | https://pgcdata.med.unc.edu/cross_disorder/pgc_cdg2_meta_no23andMe_oct2019_v2.txt.daner.txt.gz |
| Depression [2] | 264,984 | 581,929 | 122 | dbGaP Study Accession: phs001672.v6.p1 |
| Schizophrenia [3] | 36,989 | 113,075 | 108 | https://pgcdata.med.unc.edu/schizophrenia/ckqny.scz2snpres.gz |
| Bipolar disorder [4] | 20,352 | 31,358 | 16 | https://www.med.unc.edu/pgc/download-results/<br>File = daner_PGC_BIP32b_mds7a_0416a |
| Autism [5] | | | 2 | https://pgcdata.med.unc.edu/autism_spectrum_disorders/iPSYCH-PGC_ASD_Nov2017.gz |
| ADHD [6] | 19,099 | 34,194 | 10 | https://pgcdata.med.unc.edu/adhd/adhd_eur_jun2017.gz |
| Body mass index [7] | 806,834 | NA | 1023 | https://zenodo.org/record/1251813#.X_iGVS-l1TZ<br>File = bmi.giant-ukbb.meta-analysis.combined.23May2018.txt |
| Alzheimer's disease [8] | 71,880 | 383,378 | 25 | https://ctg.cncr.nl/documents/p1651/AD_sumstats_Jansenetal_2019sept.txt.gz |
| Rheumatoid arthritis [9]. | 14,361 | 43,923 | 48 | http://plaza.umin.ac.jp/~yokada/datasource/files/GWASMetaResults/RA_GWASmeta_European_v2.txt.gz |

**Table S1** Genome-wide association study details. Genetic variants associated trans-diagnostically with risk for 8 psychiatric disorders and cis-diagnostically with risks for each of 5 specific psychiatric / neurodevelopmental disorders, and 3 positive control disorders. Loci associated with risk were thresholded at $P < 5x10^{-8}$, then distance-based clumping was used to define independently significant loci (see **Methods**).

**Table S2** Code and data availability

| **Resource** | **Availability** |
|---|---|
| Code used to perform this analysis and generate the figures in the paper | https://github.com/maryellenlynall/psychimmgen2021 |
| Summary statistics | See **Table S1** |
| Roadmap Epigenomics datasets | http://bx.psu.edu/~yuzhang/Roadmap_ideas/trackDb_test.txt |
| BLUEPRINT datasets | |
| Soskic immune stimulation dataset (H3K27ac) | https://www.ebi.ac.uk/ega/studies/EGAS00001002749 |
| IDEAS annotations | http://bx.psu.edu/~yuzhang/Roadmap_ideas/trackDb_test.txt |
| 1000 genomes called against GRCh38 | http://ftp.1000genomes.ebi.ac.uk/vol1/ftp/data_collections/1000_genomes_project/release/20190312_biallelic_SNV_and_INDEL/ |
| CHEERS code | https://github.com/trynkaLab/CHEERS |
| Partitioned LD scores for active regulatory elements in Roadmap tissues | Generated in this analysis; available at https://doi.org/10.5281/zenodo.5153661 |
| GnomAD v2.1.1 | https://gnomad.broadinstitute.org |

**Table S3** Statistical comparison of original vs. conditional s-LDSC models by *Z*-test (see **Methods**), to accompany **Figure 1B** and **Figure S2A**

| | Original model vs. conditional model including fetal male brain (**Fig S2A**) | Original model vs. conditional model including fetal female brain (**Fig S2A**) |
|---|---|---|
| Brain Angular Gyrus | z=1.80; p=0.04 | z=2.19; p=0.01 |
| Brain Anterior Caudate | z=1.74; p=0.04 | z=2.20; p=0.01 |
| Brain Cingulate Gyrus | z=1.75; p=0.04 | z=2.19; p=0.01 |
| Brain Germinal Matrix | z=2.61; p=0.005 | z=2.92; p=0.002 |
| Brain Hippocampus Middle | z=1.50; p=0.07 | z=1.99; p=0.02 |
| Brain Inferior Temporal Lobe | z=1.80; p=0.04 | z=2.19; p=0.01 |
| Brain Dorsolateral Prefrontal Cortex | z=1.94; p=0.03 | z=2.31; p=0.01 |
| Brain Substantia Nigra | z=1.34; p=0.09 | z=1.77; p=0.04 |
| Fetal Brain (female) | z=1.73; p=0.04 | NA |
| Fetal Brain (male) | NA | z=2.77; p=0.003 |
| Immune cell subsets for which trans-risk showed significant enrichment (q < 0.05) in original s-LDSC model | Original model vs. conditional model including all 10 significantly enriched brain regions (**Fig 1B**) | |
| T cytotoxic naive cells (peripheral blood) | z=0.34; p=0.37 | |
| T helper naive cells (peripheral blood) 2 | z=0.20; p=0.42 | |
| T cytotoxic memory cells (peripheral blood) | z=0.21; p=0.42 | |
| T helper 17 cells (PMA-I stimulated) | z=0.19; p=0.42 | |
| T helper cells (PMA-I stimulated) | z=0.19; p=0.42 | |
| T helper memory cells (peripheral blood) 1 | z=0.17; p=0.43 | |
| T regulatory cells (peripheral blood) | z=0.20; p=0.42 | |
| T effector/memory (peripheral blood) | z=0.26; p=0.40 | |
| T helper cells (peripheral blood) | z=0.20; p=0.42 | |
| T helper naive cells (peripheral blood) 1 | z=0.28; p=0.39 | |
| T helper memory cells (peripheral blood) 2 | z=0.18; p=0.43 | |
| T cells (cord blood) | z=0.23; p=0.41 | |

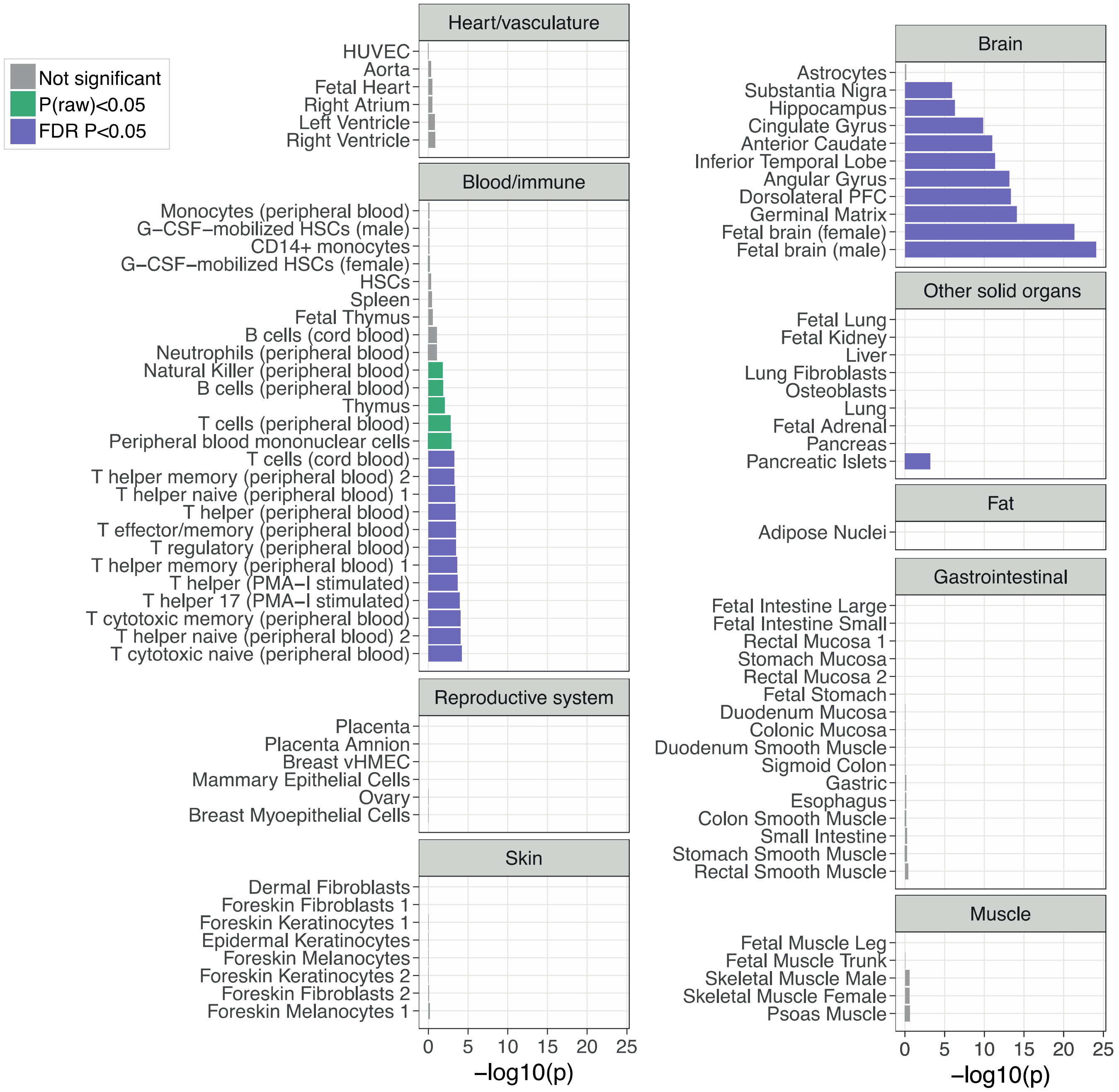


**Figure S1 Enrichment of trans-diagnostic risk at active regulatory elements (active promoters and enhancers) in 88 tissues from the Roadmap epigenomics consortium**. *P*-values are shown for the results of stratified linkage disequilibrium score regression (s-LDSC) analysis, taking the union of active elements in a given cell type as the annotation of interest (see **Methods**). The *P*-values from s-LDSC were used to test the null hypotheses that risk variants were not co-located with epigenetically activated sites more frequently than expected by chance, at two probability thresholds: $P < 0.05$ (green); and Benjamini-Hochberg $P_{FDR} < 0.05$ (purple), to correct for multiple tests across N=88 tissues. HUVEC, human umbilical vein endothelial cells; vHMEC, variant human mammary epithelial cells; PFC, prefrontal cortex; HSC, hematopoietic stem cell; PMA-I, phorbol-myristate-acetate and ionomycin. A subset of these results is shown in **Figure 1A**.

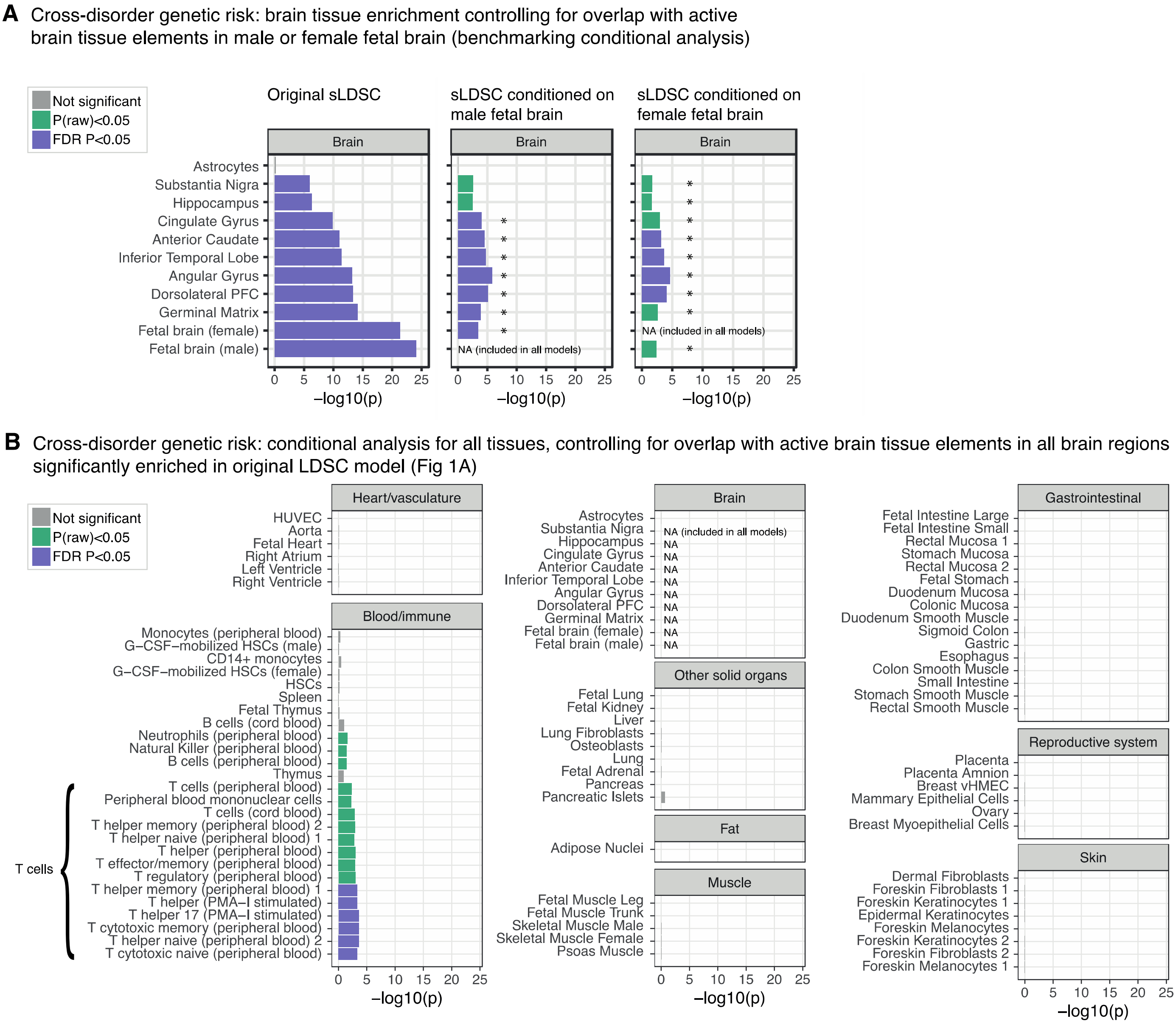


**Figure S2 Cross-disorder psychiatric risk is enriched in brain and lymphoid immune cell active genomic elements** (A) Validation of brain-conditioned analysis method: repeat of the analysis in **Figure 1A**, including the active regulatory annotation for fetal male brain as an additional term in the s-LDSC models for other cell types. LHS shows original s-LDSC for brain regions; RHS shows s-LDSC analysis with the addition of fetal male brain annotation to all models. Asterisks indicate those annotations showing significantly decreased enrichment in the conditional compared to the original analysis (two-sample $Z$-test $P < 0.05$). (B) Brain-conditioned analysis: repeat of the analysis in **Figure 1A**, including the active regulatory annotations for all 10 significantly enriched brain regions as additional terms in the s-LDSC models for all other cell types. A subset of these results is shown in **Figure 1B**.

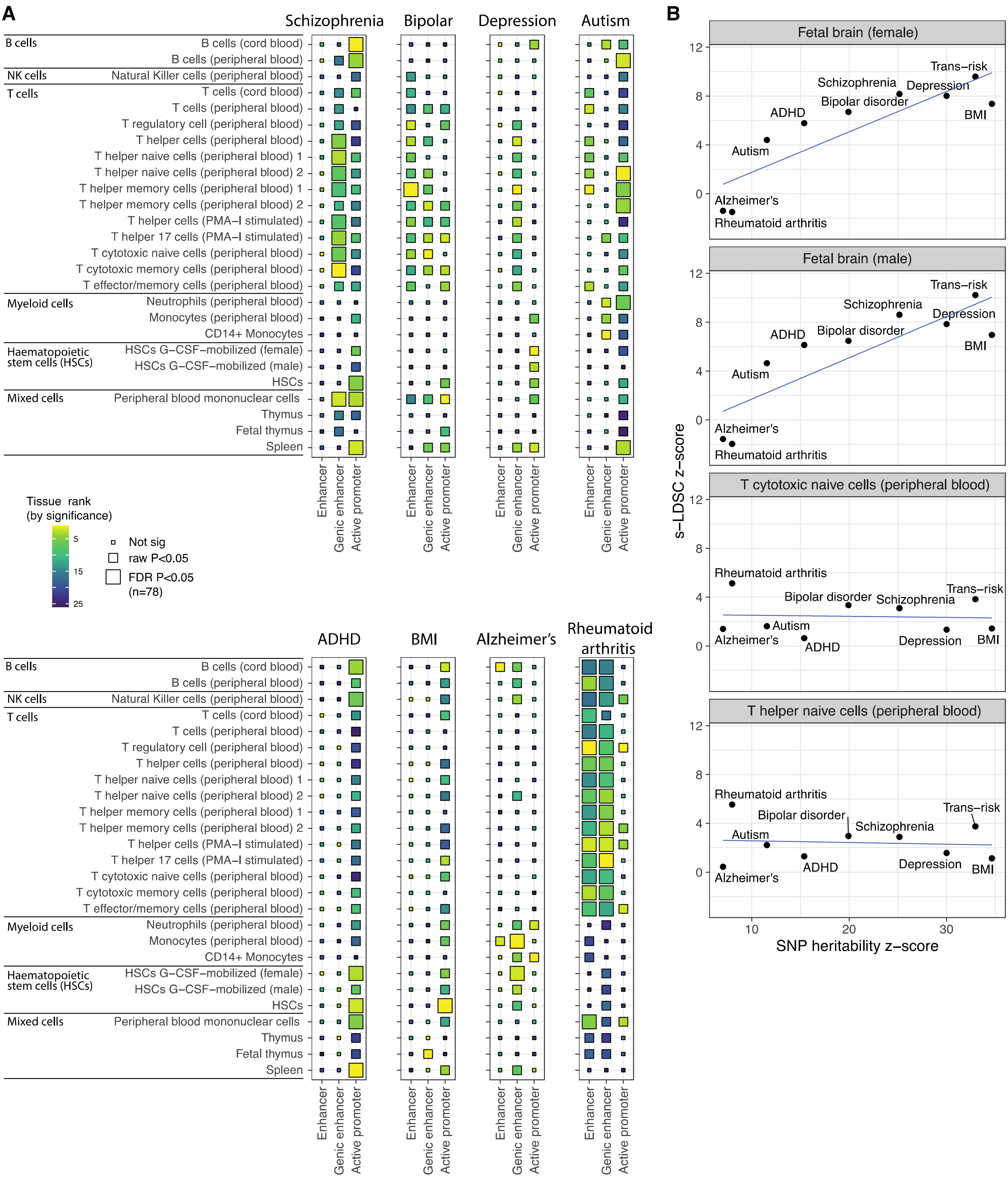


**Figure S3 Enrichment of genetic risk for multiple psychiatric disorders in immune tissues**
(A) Enrichment at immune cell enhancers, genic enhancers and active promoters in Roadmap immune tissues. Enrichment at each annotation was calculated using stratified linkage disequilibrium score regression (s-LDSC). The *P*-values from s-LDSC indicate the significance of the coefficient for the cell type specific annotations. Tile size indicates significance with FDR correction across all 78 annotations tested. Tile fill indicates the *P*-value rank (ranking within each

annotation across cell types). (B) Correlations between GWAS SNP heritability *Z*-scores and s-LDSC *Z*-scores for the top two brain and immune annotations for trans-risk. Spearman's correlations with heritability z-score are as follows: fetal male brain $\rho = 0.87$, $P = 0.005$; fetal female brain $\rho = 0.87$ $P = 0.005$; cytotoxic T cells $\rho = 0$, $P = 1$; helper T cells: $\rho = 0.03$, $P = 0.9$.

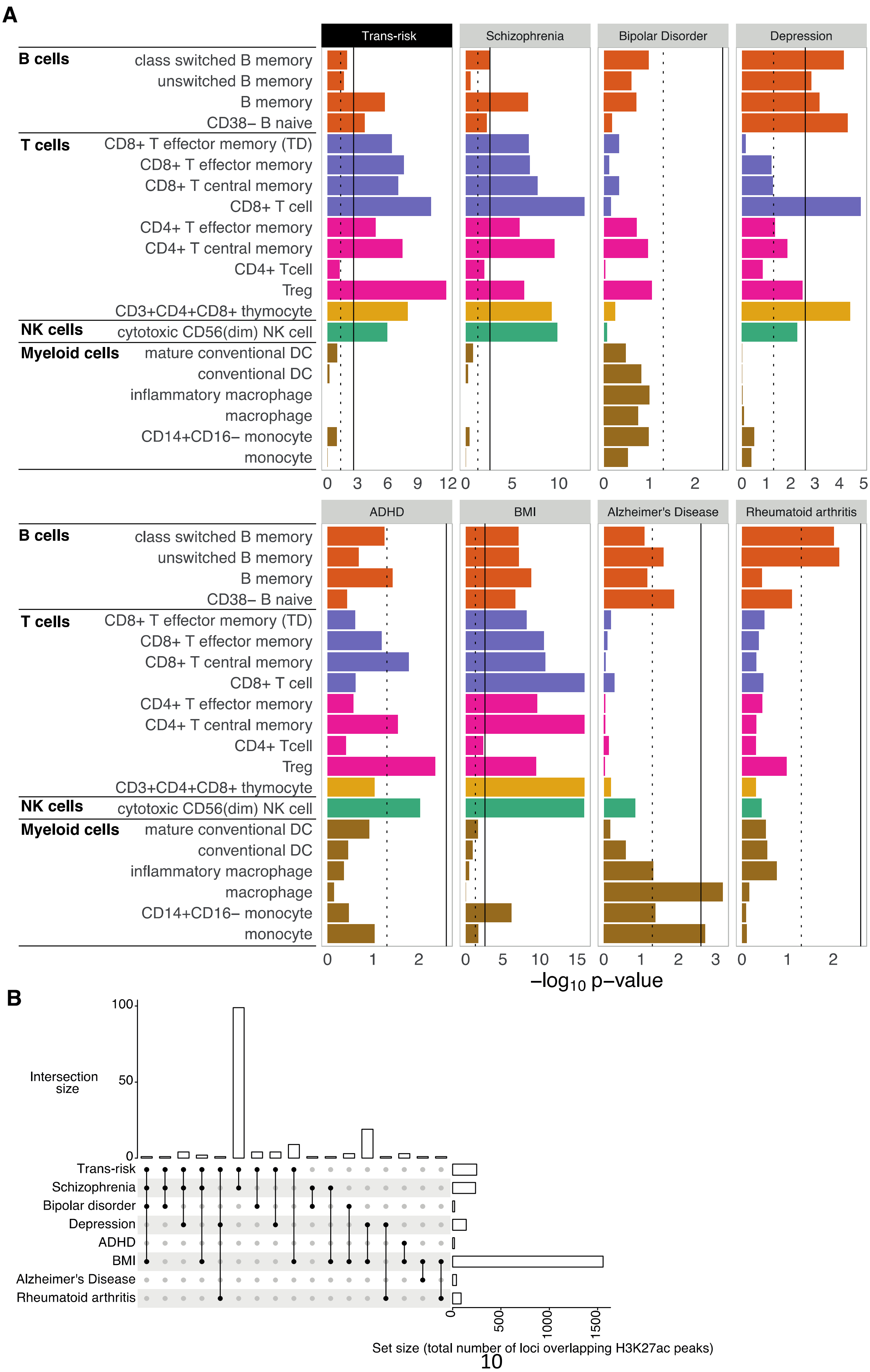
A
Trans-risk
Schizophrenia
Bipolar Disorder
Depression
ADHD
BMI
Alzheimer's Disease
Rheumatoid arthritis
B cells
class switched B memory
unswitched B memory
B memory
CD38– B naive
T cells
CD8+ T effector memory (TD)
CD8+ T effector memory
CD8+ T central memory
CD8+ T cell
CD4+ T effector memory
CD4+ T central memory
CD4+ Tcell
Treg
CD3+CD4+CD8+ thymocyte
NK cells
cytotoxic CD56(dim) NK cell
Myeloid cells
mature conventional DC
conventional DC
inflammatory macrophage
macrophage
CD14+CD16– monocyte
monocyte
–log10 p–value
B
Intersection size
Trans-risk
Schizophrenia
Bipolar disorder
Depression
ADHD
BMI
Alzheimer's Disease
Rheumatoid arthritis
Set size (total number of loci overlapping H3K27ac peaks)

113

**Figure S4 Psychiatric genetic risk enrichment at active lymphoid enhancers/promoters (BLUEPRINT dataset)** (A) Bar plots show enrichment of genetic risk for each condition at active promoters/enhancers (H3K27ac marks) in unstimulated sorted immune cells. CHEERS was used to detect enrichment of risk loci at cell-type specific H3K27ac peaks by quantifying (for each cell type) the mean cell type specificity score (for that cell type) of peaks overlapping genetic risk variants (see **Methods**). The dotted black line marks the nominal significance threshold, $P < 0.05$; the solid black line marks the Bonferroni-corrected significance threshold, $P_{FDR} < 0.05$. Note differing x-axis scales. (B) Upset plot for all Soskic immune stimulation H3K27ac immune peaks overlapped by risk variants for each disorder, showing counts (vertical bars) of shared peaks, compared to total peak number implicated by each disorder (horizontal bars).

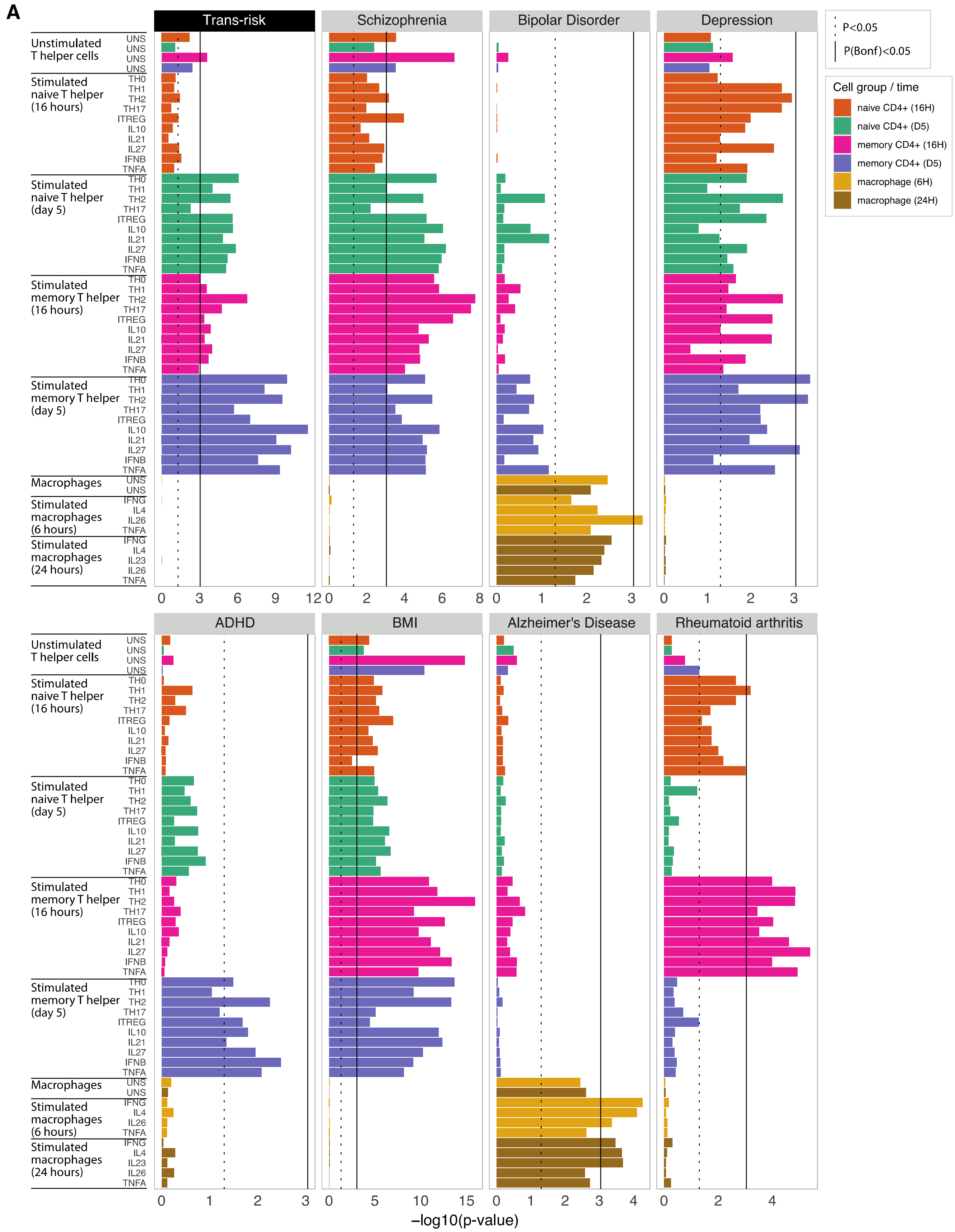


**Figure S5 Psychiatric genetic risk enrichment at activation-dependent T cell enhancers/promoters (Soskic immune stimulation dataset)**. Bar plots show enrichment of genetic risk for each condition at active promoters/enhancers (H3K27ac marks) in unstimulated and *ex vivo* stimulated immune cells. Stimulated cells are sorted macrophages, naïve $CD4^{+}$

(helper) T cells and memory $CD4^+$ T cells, assayed at both early and late timepoints (see legend). CHEERS was used to detect enrichment of risk loci at cell-type specific H3K27ac peaks by quantifying, for each cell type, the mean cell type specificity score (for that cell type) of peaks overlapping genetic risk variants (see **Methods**). P-values are reported from a discrete uniform distribution. The dotted black line marks raw $P < 0.05$; the solid black line marks Bonferroni-corrected $P_{FDR} < 0.05$. Note differing x-axis scales.

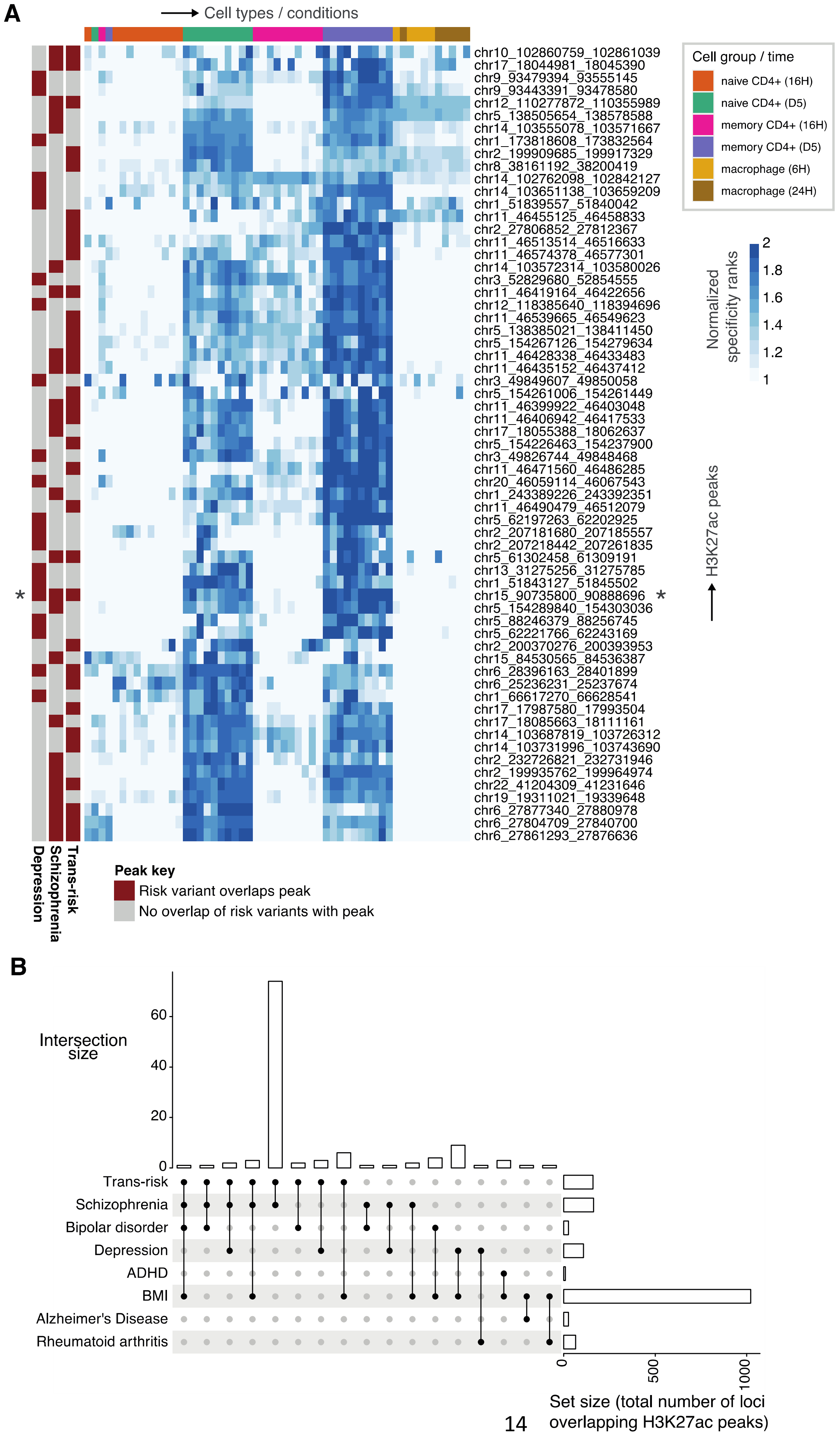
A
Cell types / conditions
chr10_102860759_102861039
chr17_18044981_18045390
chr9_93479394_93555145
chr9_93443391_93478580
chr12_110277872_110355989
chr5_138505654_138578588
chr14_103555078_103571667
chr1_173818608_173832564
chr2_199909685_199917329
chr8_38161192_38200419
chr14_102762098_102842127
chr14_103651138_103659209
chr1_51839557_51840042
chr11_46455125_46458833
chr2_27806852_27812367
chr11_46513514_46516633
chr11_46574378_46577301
chr14_103572314_103580026
chr3_52829680_52854555
chr11_46419164_46422656
chr12_118385640_118394696
chr11_46539665_46549623
chr5_138385021_138411450
chr5_154267126_154279634
chr11_46428338_46433483
chr11_46435152_46437412
chr3_49849607_49850058
chr5_154261006_154261449
chr11_46399922_46403048
chr11_46406942_46417533
chr17_18055388_18062637
chr5_154226463_154237900
chr3_49826744_49848468
chr11_46471560_46486285
chr20_46059114_46067543
chr1_243389226_243392351
chr11_46490479_46512079
chr5_62197263_62202925
chr2_207181680_207185557
chr2_207218442_207261835
chr5_61302458_61309191
chr13_31275256_31275785
chr1_51843127_51845502
chr15_90735800_90888696 *
chr5_154289840_154303036
chr5_88246379_88256745
chr5_62221766_62243169
chr2_200370276_200393953
chr15_84530565_84536387
chr6_28396163_28401899
chr6_25236231_25237674
chr1_66617270_66628541
chr17_17987580_17993504
chr17_18085663_18111161
chr14_103687819_103726312
chr14_103731996_103743690
chr2_232726821_232731946
chr2_199935762_199964974
chr22_41204309_41231646
chr19_19311021_19339648
chr6_27877340_27880978
chr6_27804709_27840700
chr6_27861293_27876636
*
Cell group / time
naive CD4+ (16H)
naive CD4+ (D5)
memory CD4+ (16H)
memory CD4+ (D5)
macrophage (6H)
macrophage (24H)
Normalized specificity ranks
2
1.8
1.6
1.4
1.2
1
H3K27ac peaks
Depression
Schizophrenia
Trans-risk
Peak key
Risk variant overlaps peak
No overlap of risk variants with peak
B
Intersection size
60
40
20
0
Trans-risk
Schizophrenia
Bipolar disorder
Depression
ADHD
BMI
Alzheimer's Disease
Rheumatoid arthritis
0
500
1000
Set size (total number of loci overlapping H3K27ac peaks)

**Figure S6 Soskic stimulated immune cell dataset: overlap of H3K27ac peaks implicated by different disorders** (A) Heatmap shows the subset of peaks with specificity for late-activated naïve and/or memory $CD4^+$ T cells which are also overlapped by risk variants for either trans-risk, schizophrenia, or major depressive disorder. Each row corresponds to a H3K27ac peak overlapping a risk variant; each column corresponds to a different cytokine-induced cell state (see legend), ordered as in **Figure 4A**. Blue fill shade represents how specific each peak is to each cell state (specificity rank of the peak normalized to the mean specificity rank of all peaks). Row annotations indicate peaks which overlap (dark red) or do not overlap (grey) risk variants for the disorder indicated. Of the late-activation T cell specific peaks, only 1 (starred *) is overlapped by both schizophrenia and depression risk variants. (B) Upset plot for all Soskic dataset H3K27ac immune peaks overlapped by risk variants for each disorder, showing counts (vertical bars) of shared peak overlaps, compared to total number of peaks implicated by each disorder (horizontal bars).

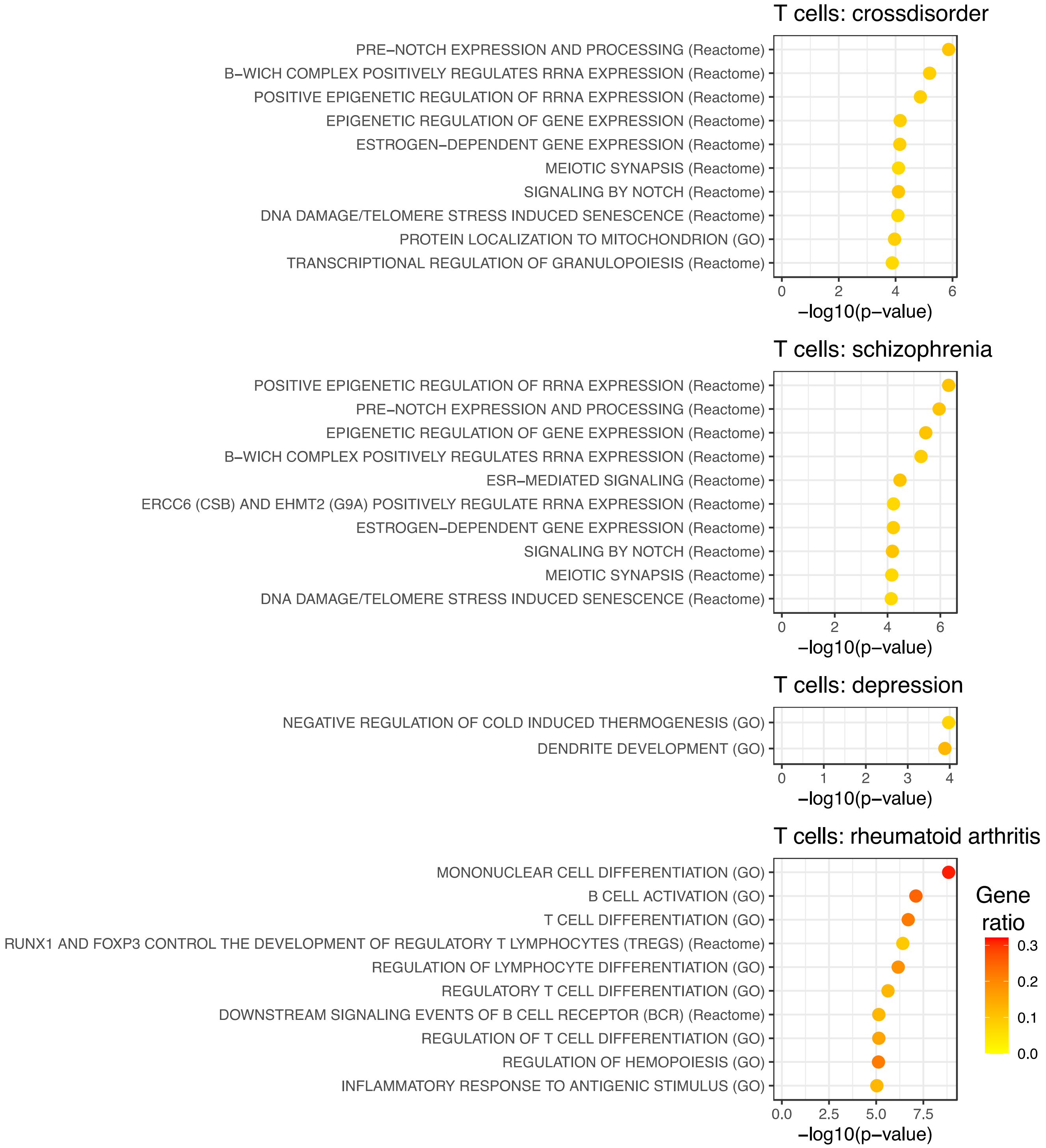


**Figure S7 Soskic stimulated immune cell dataset: pathway enrichment for genes nearest to peaks both specific to T cells and overlapped by risk variants.** Results are shown for those disorders which showed enrichment in T cell subset. Peaks which both overlapped a disease-specific risk variant and were highly specific (specificity rank > 0.9) to any of the T cell subsets in the Soskic immune stimulation dataset were selected, and nearest genes identified (see **Methods**). For these genes, over-representation analysis was used to detect pathway enrichment in Reactome and GO Biological Process pathways. Only pathways with $P_{FDR}$ < 0.05 are shown, with a maximum of 10 pathways shown per condition. Fill colour indicates gene ratio (number of test genes in the pathway / total number of test genes).
